# Multiscale Neural Modeling of Resting-state fMRI Reveals Executive-Limbic Malfunction as a Core Mechanism in Major Depressive Disorder

**DOI:** 10.1101/2020.04.29.20084855

**Authors:** Guoshi Li, Yujie Liu, Yanting Zheng, Ye Wu, Danian Li, Xinyu Liang, Yaoping Chen, Ying Cui, Pew-Thian Yap, Shijun Qiu, Han Zhang, Dinggang Shen

## Abstract

Computational neuroimaging has played a central role in characterizing functional abnormalities in major depressive disorder (MDD). However, most of existing non-invasive analysis tools based on functional magnetic resonance imaging (fMRI) are largely descriptive and superficial, thus cannot offer a deep mechanistic understanding of neural circuit dysfunction in MDD. To overcome this limitation, we significantly improved a previously developed *Multiscale Neural Model Inversion (MNMI*) framework that can link *mesoscopic* neural interaction with *macroscale* network dynamics and enable the estimation of both intra-regional and inter-regional effective connectivity. We applied the improved MNMI approach to test two well-established competing hypotheses of MDD pathophysiology (*default mode-salience* network disruption vs. *executive-limbic* network malfunction) based on resting-state fMRI with a relatively large sample size. Results indicate that MDD is primarily characterized by aberrant circuit interactions within the executive-limbic network, rather than the default mode-salience network. Specifically, we observed reduced frontoparietal effective connectivity that potentially contributes to hypoactivity in the dorsolateral prefrontal cortex (dlPFC), and decreased intrinsic inhibition combined with increased excitation from the superior parietal cortex (SPC) that potentially leads to amygdala hyperactivity, together resulting in connectivity imbalance in the PFC-amygdala circuit that pervades in MDD. Moreover, the model unravels reduced PFC-to-hippocampus excitation but decreased SPC-to-thalamus inhibition in MDD population that potentially leads to hypoactivity in hippocampus and hyperactivity in thalamus, consistent with previous experimental data. Overall, our findings provide strong support for the long-standing limbic-cortical dysregulation model in major depression but also offer novel insights into the multi-scale pathophysiology of this deliberating disease.

## Introduction

Major depressive disorder (MDD) is a serious mental illness that is characterized by depressed mood, diminished interests and impaired cognitive function (Otte et al., 2016). It is a leading cause of chronic disability worldwide with a lifetime prevalence of up to 17% (Kessler et al., 2005). Despite the advances in treatment modalities, it is estimated that up to 30% of patients do not respond to standard intervention (Raymaekers et al., 2017) and 10%–30% develop treatment-resistant symptoms with increased disability and suicidal tendency as well as a higher risk of relapse (Al-Harbi, 2012; Delaloye and Holtzheimer, 2014). To improve diagnosis and treatment of MDD, a better understanding of the pathophysiological mechanisms of MDD is urgently needed.

Although the exact mechanisms underlying MDD remain unclear, clinical, neuroimaging and animal studies are starting to offer promising insights into the neurobiological basis of this debilitating disease. Preclinical stress models in rodents and non-human primates have reported disrupted excitatory and inhibitory neurotransmission along with neuronal atrophy in cortical and limbic regions, leading to structural and dynamical abnormalities (Musazzi et al., 2011; Duman et al., 2019). Consistently, structural magnetic resonance imaging (MRI) and functional MRI (fMRI) have revealed both structural and functional brain alterations in MDD (Otte et al., 2016; Lener et al., 2017). Understanding the impaired interactions between brain regions is of particular importance as it is generally believed that MDD can be characterized as a network disorder with aberrant connections among various brain areas within networks (Menon, 2011; Dutta et al., 2014; Mulders et al., 2015; Brakowski et al., 2017). Indeed, both task-based and resting-state fMRI (rs-fMRI) indicate disrupted connectivity among several functional networks in the large-scale brain pathophysiology of MDD (Kerestes et al., 2014; Kaiser et al., 2015; Yang et al., 2016; Drysdale et al., 2017; Zhou et al., 2019). The first highly implicated large-scale brain network is the default mode-salience network. The default model network (DMN) is activated in the resting brain and consists of ventral anterior cingulate cortex (vACC), posterior cingulate cortex (PCC)/precuneus, medial prefrontal cortex (mPFC), and lateral and inferior parietal cortex (Raichle et al., 2001; Dutta et al., 2014). The DMN has been implicated in rumination and self-referential processing (Cooney et al., 2010; Liston et al., 2014) and increased intrinsic connectivity in the DMN is hypothesized to underlie excessive self-focused rumination in MDD (Cooney et al., 2010; Hamilton et al., 2015). The salience network, on the other hand, is activated in response to salient stimuli including emotional pain, empathy, metabolic stress and social rejection (Seeley et al., 2007; Hermans et al., 2014). Anchored in the dorsal anterior cingulate cortex (dACC) and anterior insula, the salience network is involved in integrating highly processed sensory information with visceral, autonomic and hedonic states, making it particularly important for interoceptive-autonomic processing (Seeley et al., 2007). Disruption of DMN-SAL connectivity has been reported in a number of MDD studies. Manoliu et al. (2014) found that MDD patients showed increased FC between DMN and SAL, consistent with greater resting-state FC between insula and anterior DMN (Avery et al., 2014). It has also been observed that FC between anterior DMN and SAL was positively correlated with depression severity in post-stroke depression (Balaev et al., 2017). Using dynamic causal modeling of resting-state fMRI (rs-fMRI), we recently demonstrated that MDD was mainly associated with reduced effective connectivity (EC) within the DMN, and between the DMN and SAL networks (Li et al., 2020). These neuroimaging connectome studies support the notion that aberrant mapping or engagement of the DMN by SAL results in excessive rumination and abnormal interoceptive-autonomic processing in depression (Menon, 2011).

Another highly implicated neural system in MDD is the executive-limbic network. Opposite to the DMN, the executive control network (EXE) is most active during cognitive tasks (Seeley et al., 2007) and consists mainly of the dorsolateral prefrontal cortex (dlPFC) and posterior parietal cortex (PPC) (Corbetta and Shulman, 2002; Rogers et al., 2004; Seeley et al., 2007). The executive network plays a critical role in working memory maintenance, rule-based problem solving and decision making during goal-directed cognitive tasks (Miller and Cohen, 2001; Petrides, 2005; Koechlin and Summerfield, 2007) as well as the regulation of emotional processing (Davidson et al., 2002; Phillips et al., 2008). Disruption of the executive network has been associated with major psychiatric and neurological disorders including depression, Alzheimer’s disease (AD) and schizophrenia (Menon, 2011). The limbic system, comprised of brain regions such as the amygdala, hippocampus and thalamus, underlies important brain functions including emotion, consciousness, motivation and long-term memory (LeDoux, 2000; Rolls, 2015). The involvement of the limbic system in the pathophysiology of MDD has been well documented (Drevets et al., 2008; Pandya et al., 2012; Drysdale et al., 2017) and a cognitive model places executive-limbic dysregulation in the center of MDD pathology (Disner et al., 2011). A key component of this model lies in negative cognitive bias that is mediated by increased emotional processing from limbic regions along with diminished cognitive control. Specifically, cognitive bias is believed to be initiated by limbic hyperactivity (most notably the amygdala) and spread to the prefrontal cortex (PFC) through subgenual cingulate cortex and anterior cingulate cortex (ACC). Concurrently, hypoactivity in the dlPFC reduces the cortical control over the limbic system further heightening the amygdala’s responses, which in turn reinforces the cognitive biases and leads to persistent increased awareness and conscious processing of negative stimuli (Disner et al., 2011). As a result, depressive mood and negative thoughts perpetuate in depression. The executive-limbic dysregulation model has received substantial experimental support in the literature. Individuals with depression showed much more intense (up to 70%) and longer (up to three times) response in the amygdala than healthy subjects in response to negative stimuli (Drevets, 2001; Siegle et al., 2002). It has also been observed that the amygdala response in MMD patients increased linearly as the intensity of a sad facial expression increased (Surguladze et al., 2005). By comparison, the dlPFC exhibits lower response to negative stimuli in individuals with MDD than healthy subjects (Hamilton et al., 2012; Fales et al., 2008) and hypoactivation in the dlPFC is associated with excessive rumination (Ochsner et al., 2004; Ray et al., 2005). In a cognitive control task involving attentional focus shifting, participants with elevated depressive symptoms displayed weaker activation in the lateral PFC and parietal regions compared with subjects with mild depression (Beevers et al., 2010). In addition to activation imbalance, abnormal connectivity patterns have been observed in PFC-amygdala coupling (Johnstone et al., 2007; Dannlowski et al., 2009; de Almeida et al., 2009; Lu et al., 2012).

Although the two neurobiological models of depression (default mode-salience disruption versus executive-limbic malfunction) have each received considerable experimental support, it is not known which model plays a more fundamental role in MDD pathophysiology. Addressing this question will shed light on the core neural circuitry mediating depression and the auxiliary circuitry that is likely subject to the influence of the core network. In addition, the existing connectome studies of MDD have focused predominantly on undirected correlation (i.e., FC estimated by Pearson’s correlation) of fMRI blood-oxygen-level-dependent (BOLD) signals, which neither provides the causal interaction relationship among different brain regions nor offers a neurophysiological account of the aberrant neural process. Effective connectivity (EC), by comparison, characterizes the causal influence that one neural system exerts over another, which can better explain the causal relationship of observed signals than FC (Friston, 2011). As a predominant approach to compute EC, Dynamic Causal Modeling (DCM) employs a model of neural interactions that describes distributed neuronal activity (Friston, et al., 2003), and has been applied to identify impaired inter-regional causal interactions among different brain regions in various functional networks in MDD based on either task-fMRI (Schlosser et al., 2008; de Almeida et al., 2009, 2011; Lu et al., 2012) or rs-fMRI (Hyett et al., 2015; Li et al., 2017; Kandilarova et al., 2018; Li et al., 2020). However, the underlying neural model of DCM for fMRI largely relies on a highly simplified linear state-space model for generating neural activity (Friston, et al., 2003), which cannot capture impaired neural interactions at finer cellular and circuit levels. On the other hand, recent advance in basic neuroscience and high performance computing has enabled the development of large-scale biologically realistic or plausible neuronal models to study neural circuit functions (e.g., Hoang et al., 2013; Markram et al., 2015; Schmidt et al., 2018). A better integration of computational neuroscience with connectome modeling is a crucial next step to advance our understanding of the pathophysiological mechanisms of MDD.

To overcome the aforementioned limitations of traditional EC studies, we have previously developed a Multiscale Neural Model Inversion (MNMI) framework that can link *mesoscale* intra-regional circuit interactions with *macroscale* inter-network dynamics and enable the estimation of both intra-regional and inter-network EC based on rs-fMRI (Li et al., 2019). Nevertheless, as a preliminary study, we estimated the EC among different functional networks instead of different brain regions due to the scale limit (i.e., the inter-regional EC was restricted to inter-network EC to reduce the freely estimable parameters), which may preclude the detection of more subtle EC change at single region level (i.e., between a pair of regions). In addition, the inter-network EC was assumed to be positive (excitatory), which cannot account for the potentially inhibitory effect between certain brain regions (e.g., the inhibitory effect from PFC to amygdala, Quirk et al., 2003). Here, we refined the MNMI framework by not only enabling inter-regional EC estimation but also relaxing the positive constrain on inter-regional EC. Notably, we came up with a structural connectivity-based method to reduce the number of inter-regional connections to those with strong structural connectivity in order to avoid the problem of over-fitting and improve estimation efficiency for more comprehensive and realistic modeling. By applying the refined MNMI approach to a relatively large rs-fMRI dataset consisting of 98 healthy normal control (NC) subjects and 96 individuals with first-episode drug-naïve (FEDN) MDD, we tested whether FEDN MDD was better characterized by default mode-salience or executive-limbic disruption. Results indicated that FEDN MDD was primarily characterized by abnormal circuit interactions in the executive-limbic network. Specifically, we found impaired frontoparietal interactions that could potentially lead to dlPFC hypoactivity and abnormal intrinsic inhibition combined with disrupted cortical-limbic projection that may underline amygdala hyperactivity, together resulting in PFC-amygdala activation imbalance that contributes to MDD symptoms. Overall, this computational modeling study suggests that executive-limbic malfunction constitutes the core mechanism of MDD that affects other brain functional networks and disrupted circuit interactions can be manifested during the first episode of depression enabling early diagnosis and intervention.

## Results

### Overview of the MNMI framework

The schematic diagram of the MNMI framework is depicted in Fig. 1. Each brain region consists of one excitatory and one inhibitory neural population whose intrinsic dynamics is modeled using the biologically plausible Wilson-Cowan oscillators (Wilson and Cowan, 1972). The excitatory neural population excite the inhibitory neural population and receive feedback inhibition from the inhibitory population as well as recurrent excitation from itself. The excitatory neural populations within different brain regions are interconnected via long-range structural connectivity links consisting of white-matter fibers whose baseline connection strength is determined by diffusion tractography from diffusion MRI. The simulated neural activity is converted to simulated rs-fMRI BOLD signals through a biophysical hemodynamic model (Friston et al., 2003) and the corresponding FC matrix is constructed. Then, genetic algorithm, a biologically inspired optimization algorithm, is applied to estimate both local (intra-regional) and long-range (inter-regional) ECs in order to minimize the difference between the simulated and empirical FC matrices (see Materials and Methods). Compared to the previous version of the MNMI framework (Li et al., 2019), the major changes include: 1) Inter-regional instead of inter-network ECs are separately estimated; 2) Inter-regional EC can be either positive (excitatory) or negative (inhibitory); 3) The optimization algorithm is changed from a less efficient Expectation-Maximization algorithm to Genetic Algorithm, with the latter allowing to achieve both 1) and 4), a further expanded simulation length of the neural/BOLD activity (from 80 s to 200 s); and 5) a structural connectivity-guided inter-regional EC estimation where only strong inter-regional connections are allowed in the model.

**Figure 1.**
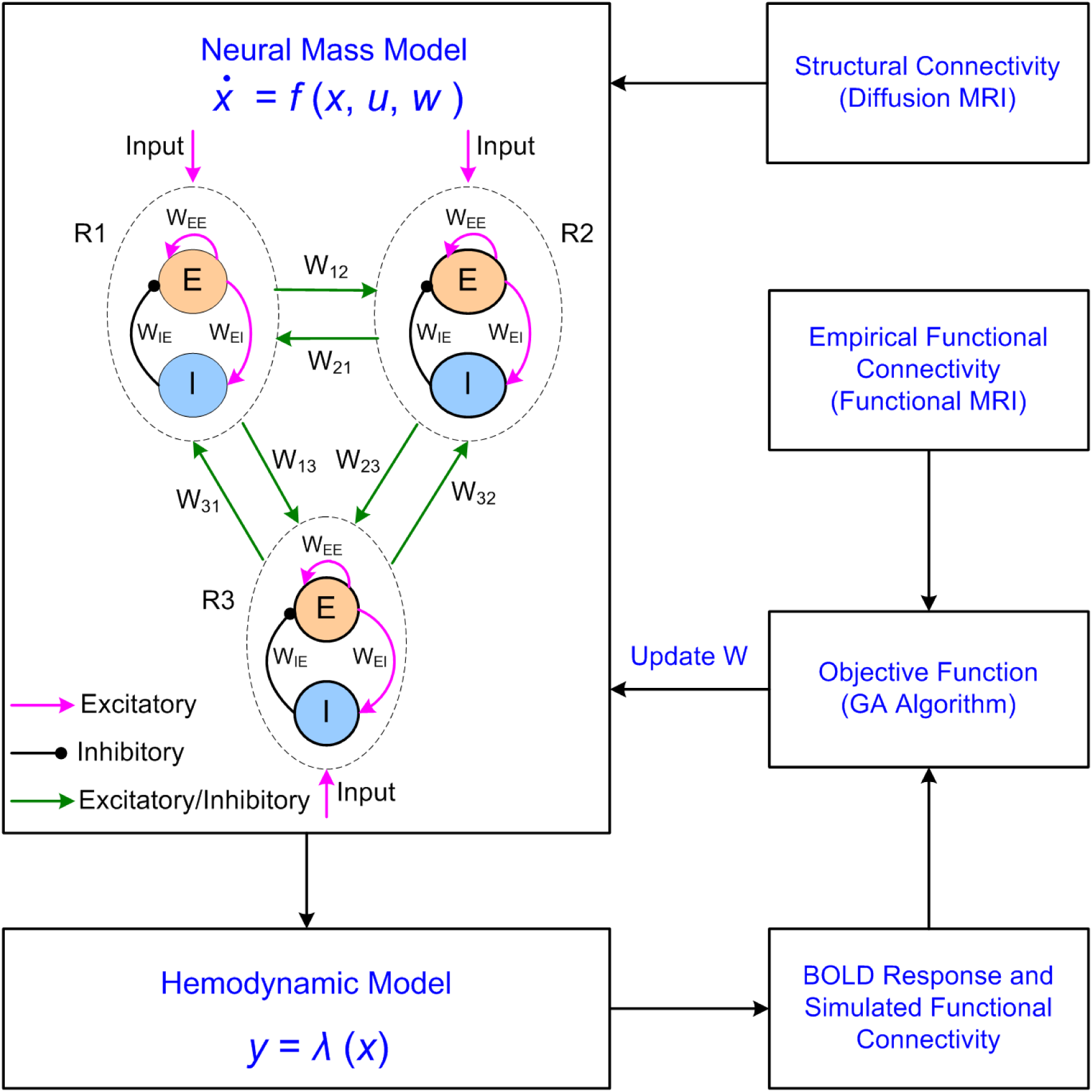
A multiscale neural model inversion (MNMI) framework to identify both intra-regional and inter-regional effective connectivity based on diffusion MRI and resting-state fMRI data. The neural activity is generated by a neural mass model consisting of multiple brain regions (R1, R2, R3, etc). Each brain region contains one excitatory (E) and one inhibitory (I) neural populations coupled with reciprocal local synapses. The excitatory neural population excite the inhibitory neural population while receiving feedback inhibition from the latter as well as recurrent excitation from itself. Different brain regions are connected via long-range fibers whose baseline connection strength is determined by structural connectivity based on diffusion MRI. The neural activity is converted into corresponding BOLD signal via a biophysical hemodynamic model and the simulated functional connectivity (FC) is computed. The objective function is calculated as the opposite of the Pearson’s correlation between the empirical and functional FC which is minimized by genetic algorithm (GA). The model parameters (both local and inter-regional connection strength) are updated iteratively until convergence is achieved.

### Participants

There was no significant difference in age (*p* = 0.99, two-sample t-test), gender (*p* = 0.35, chi-square test), or education (*p* = 0.34, two-sample t-test) between the NC and FEDN groups. The disease duration was 8.76 ± 11.04 months, and the HDRS-17 score was 22.08 ± 3.10 for the FEDN MDDs (Table 1).

**Table 1.** Demographic and clinical characteristics of participants

| Characteristics | FEDN (n = 96) | NC (n = 98) | *t*/$\chi^2$ | *p* |
| --- | --- | --- | --- | --- |
| Age (years) | 29.64 ± 9.51<sup>a</sup> | 29.48 ± 10.29 | -0.02 | 0.99<sup>b</sup> |
| Gender (F/M) | 63/33 | 58/40 | 0.86 | 0.35<sup>c</sup> |
| Education (years) | 12.51 ± 3.26 | 12.93 ± 2.77 | -0.96 | 0.34<sup>b</sup> |
| Duration (months) | 8.76 ± 11.04 | NA | NA | NA |
| HDRS-17 | 22.08 ± 3.10 | NA | NA | NA |

Abbreviations: FEDN, first-episode drug-naïve major depressive disorder; NC, normal control; HDRS-17, 17-item Hamilton Depression Rating Scale.
<sup>a</sup>Mean ± standard deviation.
<sup>b</sup>The *p* values were obtained by two-sample *t*-test.
<sup>c</sup>The *p* value was obtained by a chi-square test.

### Performance of genetic algorithm

The structural connectivity matrices of the default mode-salience network and executive-limbic networks derived from the diffusion MRI are shown in Fig. 2A, B, respectively. In the default mode-salience network, the strongest connections existed between vACC/PCC and dACC followed by the connections between vACC and PCC. In the executive-limbic network, relatively high level of connections appeared between left and right SPC, thalamus and SPC, thalamus and hippocampus, and amygdala and hippocampus. According to the one-sample *t*-test results, strong links were preserved (green edges in Fig. 2C, D). Of note, most of within-network connections were removed in the default mode-salience network and the discarded connections mostly involved the left dlPFC and left amygdala in the executive-limbic network. The genetic algorithm converged within 128 iterations/generations for all the subjects and within 80 generations for most subjects. Exemplary convergence curves for the default mode-salience and executive-limbic networks from a randomly selected NC subject are separately shown in Fig. 3A, B. The model converged rapidly within the first 20 generations and became relatively flat after 40 generations until convergence. The averaged final model fitness value (measured by Pearson correlation between the simulated and empirical FC matrices) was 0.92 for both NC and MDD groups without significant difference for the default mode-salience network (*p* = 0.8, two-sample *t*-test; Fig. 3C) and 0.87 for the executive-limbic network (*p* = 0.49, two-sample *t* -test; Fig. 3D). By comparison, the fitness value in the executive-limbic network was significantly lower than that in the default mode-salience network for both NC (*p* < 0.05, two-sample *t*-test) and MDD (*p* < 0.05, two-sample *t*-test) groups. Such difference largely reflects the fact that the dimension of the FC matrix in the executive-limbic network (9 × 9) was larger than that in the default mode-salience network (7 × 7). The simulated results for the default mode-salience and executive-limbic networks (using the optimized parameters) from a randomly selected NC subject are shown in Figs. 4 and 5, respectively. We found that the simulated neural activity and BOLD signals displayed rhythmic fluctuations in a fast and slow temporal scale, respectively. The oscillation frequency of the simulated neural activity was about 7 Hz, which resembled alpha oscillations during relaxed wakefulness (Hughes and Crunelli, 2005). The oscillation frequency of the simulated BOLD signals ranged between 0.02 and 0.08 Hz, consistent with previous experimental observations (Tong et al., 2019). There was no significant difference between NC and MDD groups, for both default mode-salience (NC: 0.06 ± 0.008 Hz; MDD: 0.06 ± 0.009 Hz, *p* = 0.29, two-sample t-test) and executive-limbic networks (NC: 0.05 ± 0.013 Hz; MDD: 0.05 ± 0.013 Hz, *p* = 0.74, two-sample t-test). In addition, the pattern of the simulated FC matched closely with that of the empirical FC (comparing Fig. 4C with 4D, and Fig. 5C with 5D). Overall, the genetic algorithm performed effectively in finding out the optimal parameters that generated realistic and biologically meaningful neural signals and led to mimic simulated FC matrix to the empirical one.

**Figure 2.**
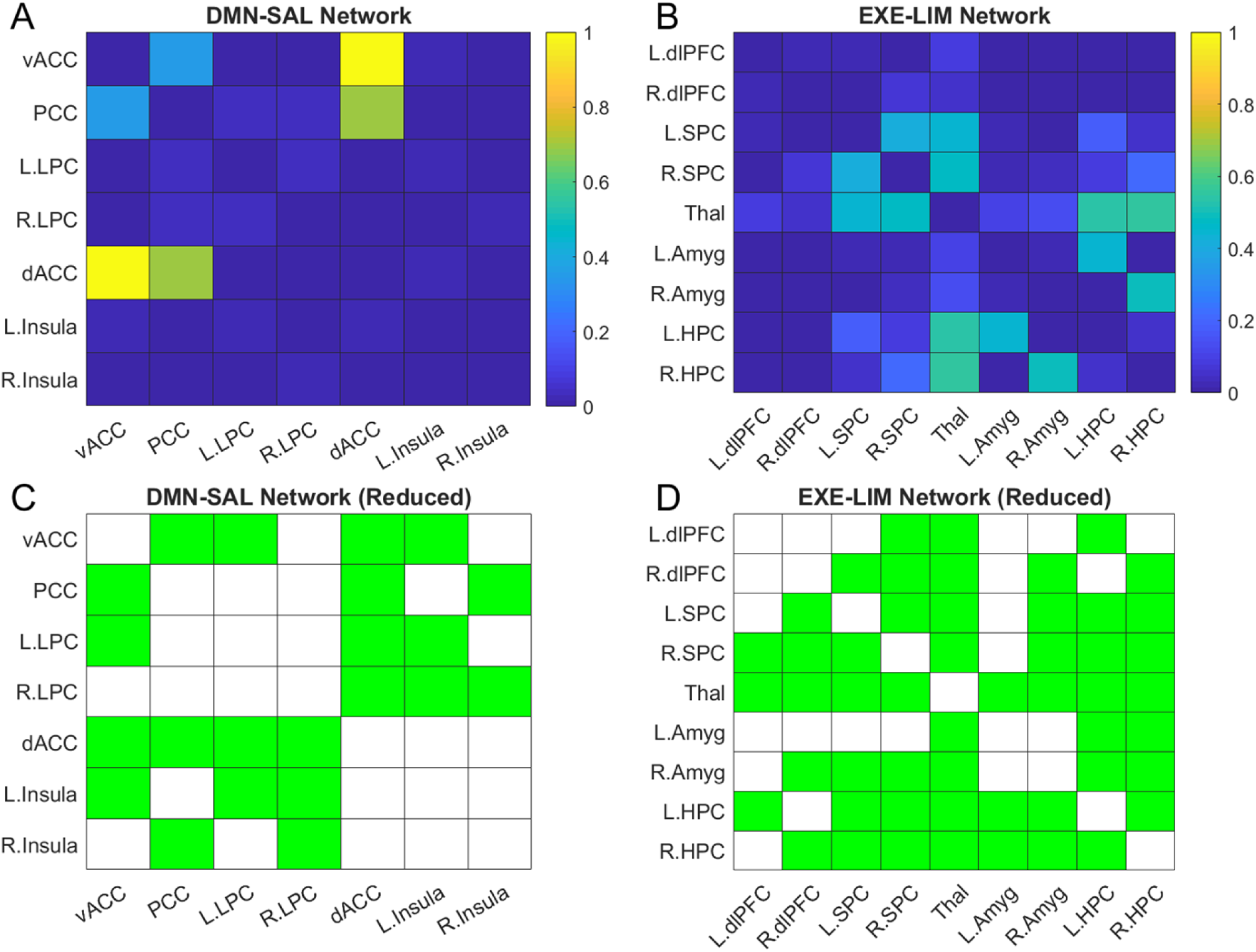
Structure connectivity of the DMN-SAL network and the EXE-LIM network with corresponding reduced networks. **(A)** Normalized structure connectivity of the DMN-SAL network. **(B)** Normalized structure connectivity of the EXE-LIM network. **(C)** Reduced DMN-SAL network; green indicate the connections that remain in the network. **(D)** Reduced EXE-LIM network; green indicate the connections that remain in the network.

**Figure 3.**
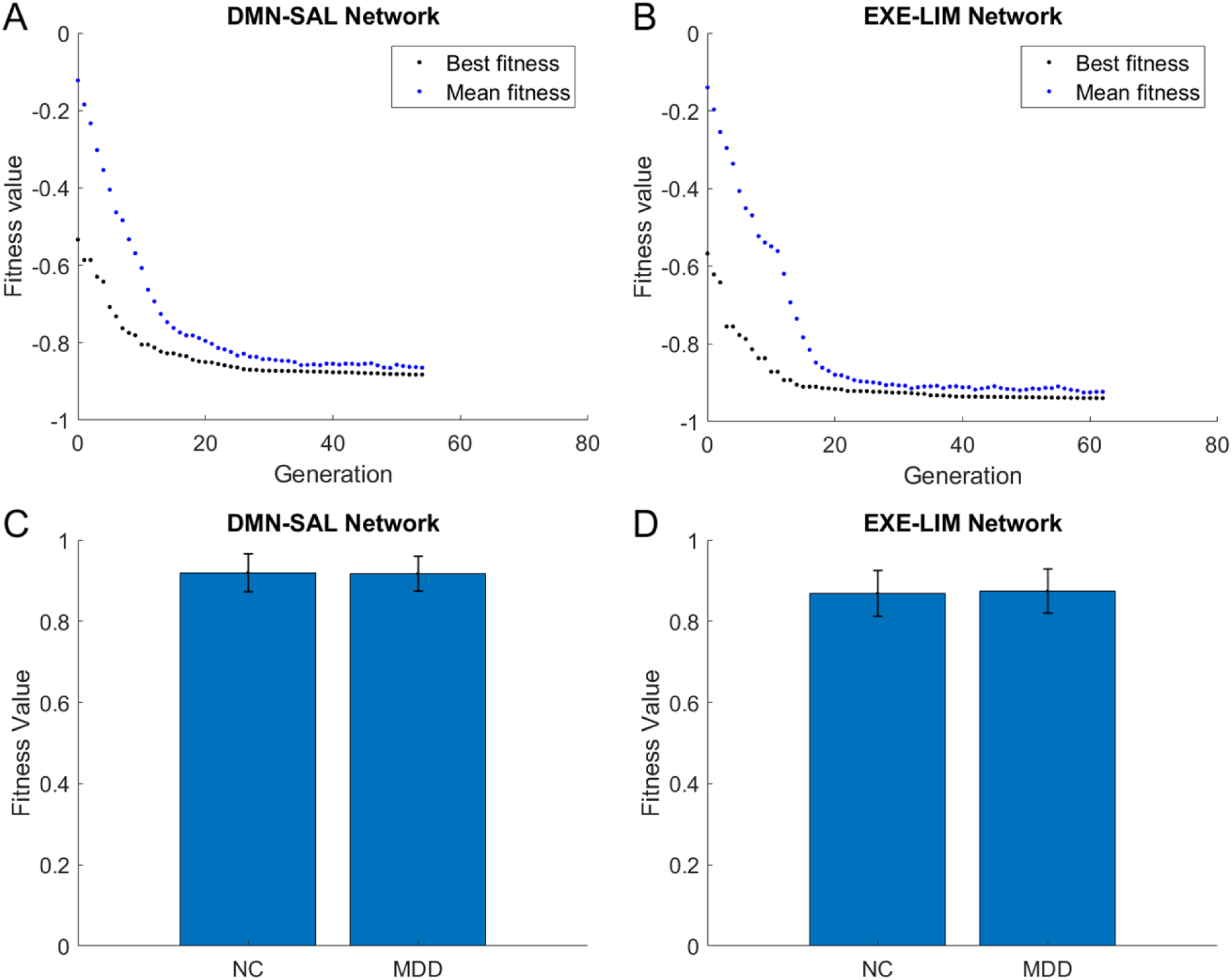
Convergence of the genetic algorithm (GA). **(A)** Convergence of the GA for the DMN-SAL network (one subject). Blue dots: mean fitness value; black dots: best fitness value. **(B)** Convergence of the GA for the EXE-LIM network (one subject). **(C)** Average fitness value for the DMN-SAL network; **(D)** Average fitness value for the EXE-LIM network. Error bars indicate standard deviation.

**Figure 4.**
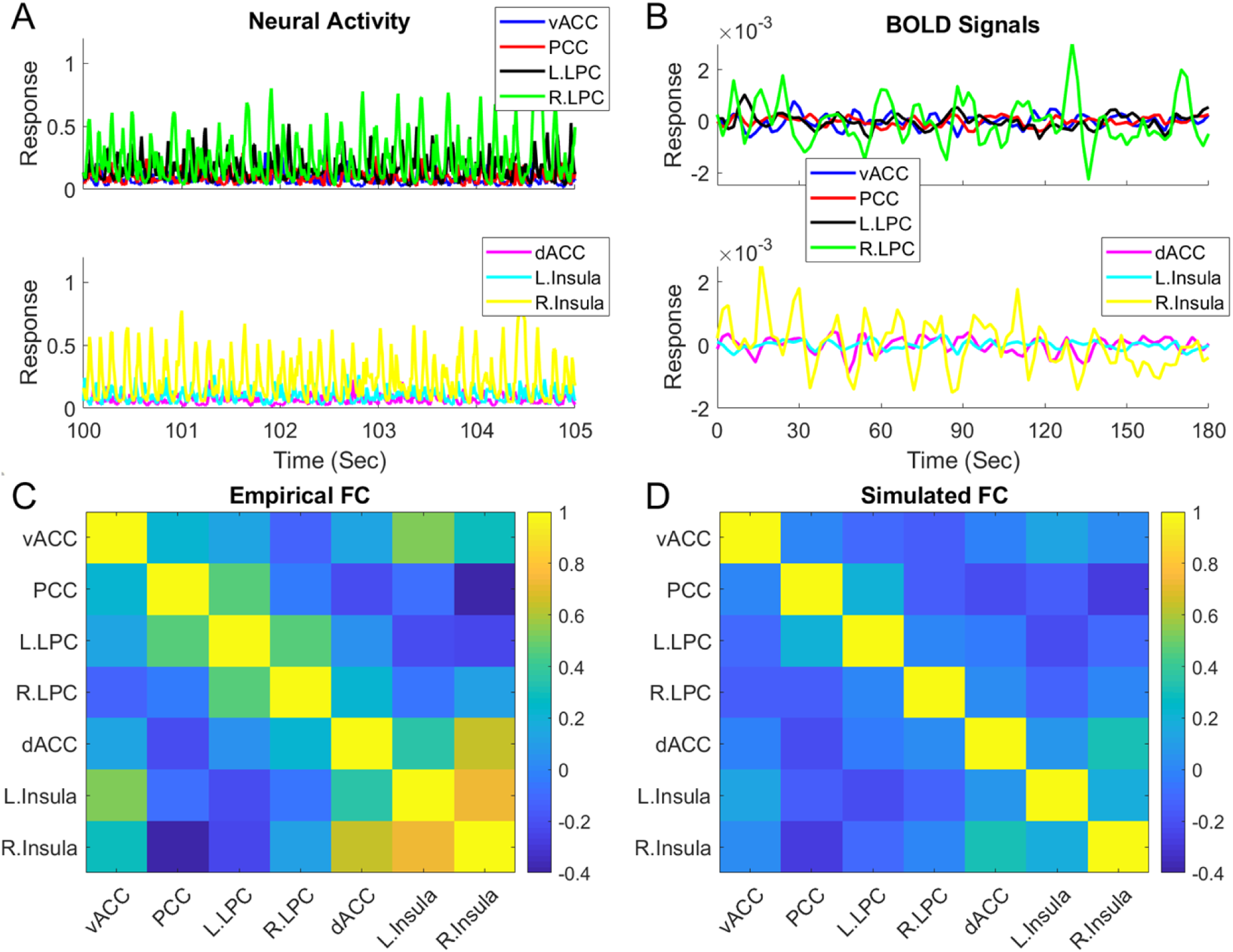
Simulated output of the DMN-SAL network for one representative subject. **(A)** A segment of simulated activity of excitatory neural populations from the seven ROIs. **(B)** Simulated BOLD signals from the seven ROIs (mean removed). **(C)** Empirical functional connectivity. **(D)** Simulated functional connectivity.

**Figure 5.**
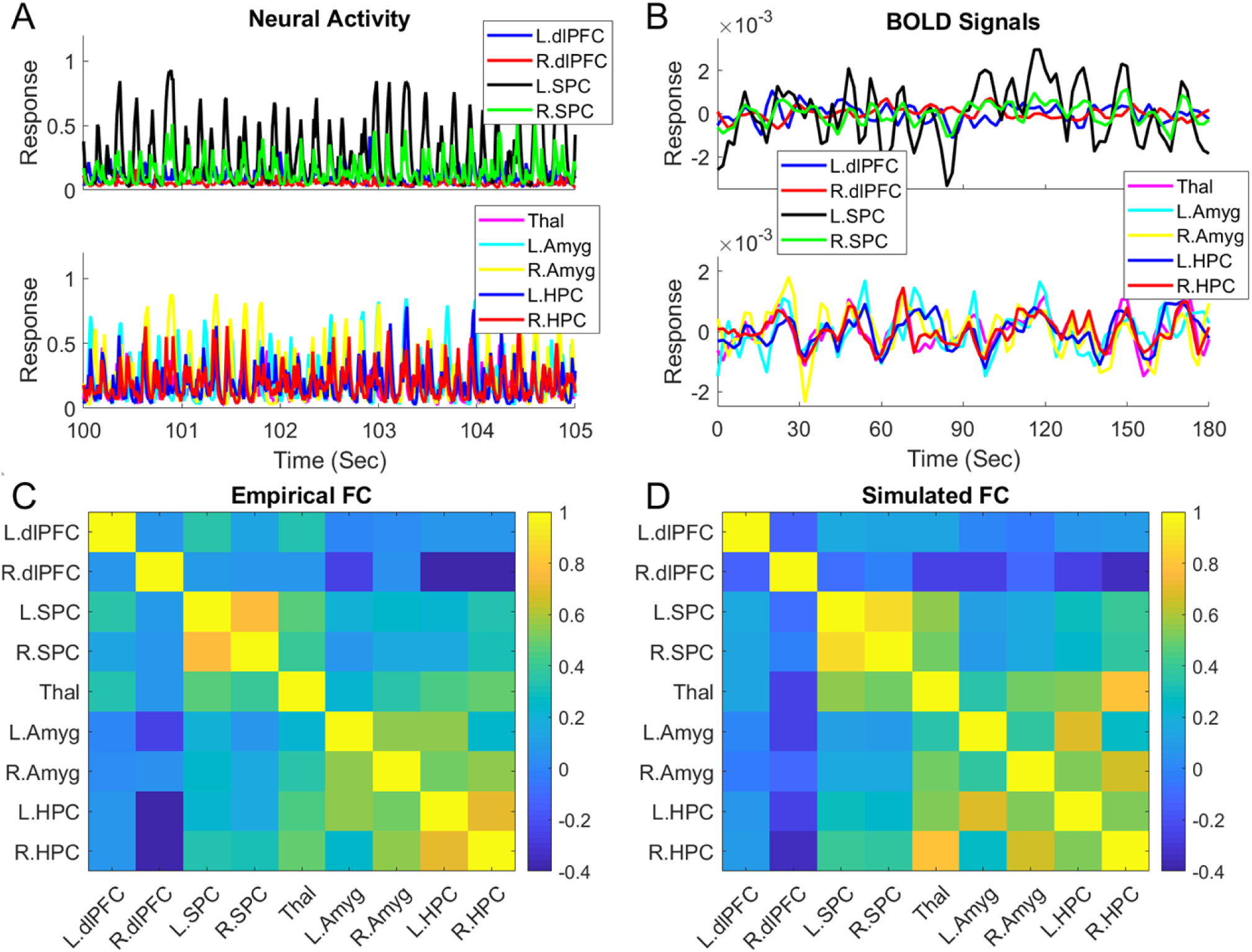
Simulated output of the EXE-LIM network for one representative subject. **(A)** A segment of simulated activity of excitatory neural populations from the nine ROIs. **(B)** Simulated BOLD signals from the nine ROIs (mean removed). **(C)** Empirical functional connectivity. **(D)** Simulated functional connectivity.

### Marginal EC group differences in the default mode-salience network between NC and MDD

The group average (and variability of) recurrent excitation and recurrent inhibition weights within each of the seven ROIs in the default mode-salience network for each group are shown in Fig. 6A, B, respectively, where no significant MDD vs. NC group difference was observed. The network-averaged recurrent excitation and inhibition weights within the DMN and SAL network are shown in Fig. 6C, D respectively. Again, there was no significant difference between the NC and MDD groups. Notably, the average recurrent excitation weight in the DMN was slightly higher than that in the SAL, while the average recurrent inhibition weight was slightly lower than that in the SAL, for both NC and MDD groups. The relatively high excitation with low inhibition may underlie the higher activation of the DMN during the resting state. The average inter-regional coupling weight within the default mode-salience network is shown in Fig. 7. Out of the 22 estimated inter-regional weights, only the vACC➜dACC connection strength was significantly (*p* < 0.05, uncorrected) decreased from -0.43 (for NCs) to -0.07 (for MDDs); but such a result was not significant after correction for multiple comparisons (which may only indicate marginal difference). To ensure that the lack of significant results was not due to removal of certain inter-regional connections, we applied the MNMI approach to the full default mode-salience network and estimated all 42 inter-regional EC links (not conducting the structural connectivity-based pre-selection but excluding self-connection weights) with results shown in Supplemental figure S5 (no significant difference in any local connection weight) and figure S6 (no significant difference in any inter-regional EC weight after multiple correction). Therefore, the model indicated that in the default mode-salience network, the intra-regional connection weight may not differ between NCs and MDDs, while the inter-regional EC may have marginal difference in one or two connections.

**Figure 6.**
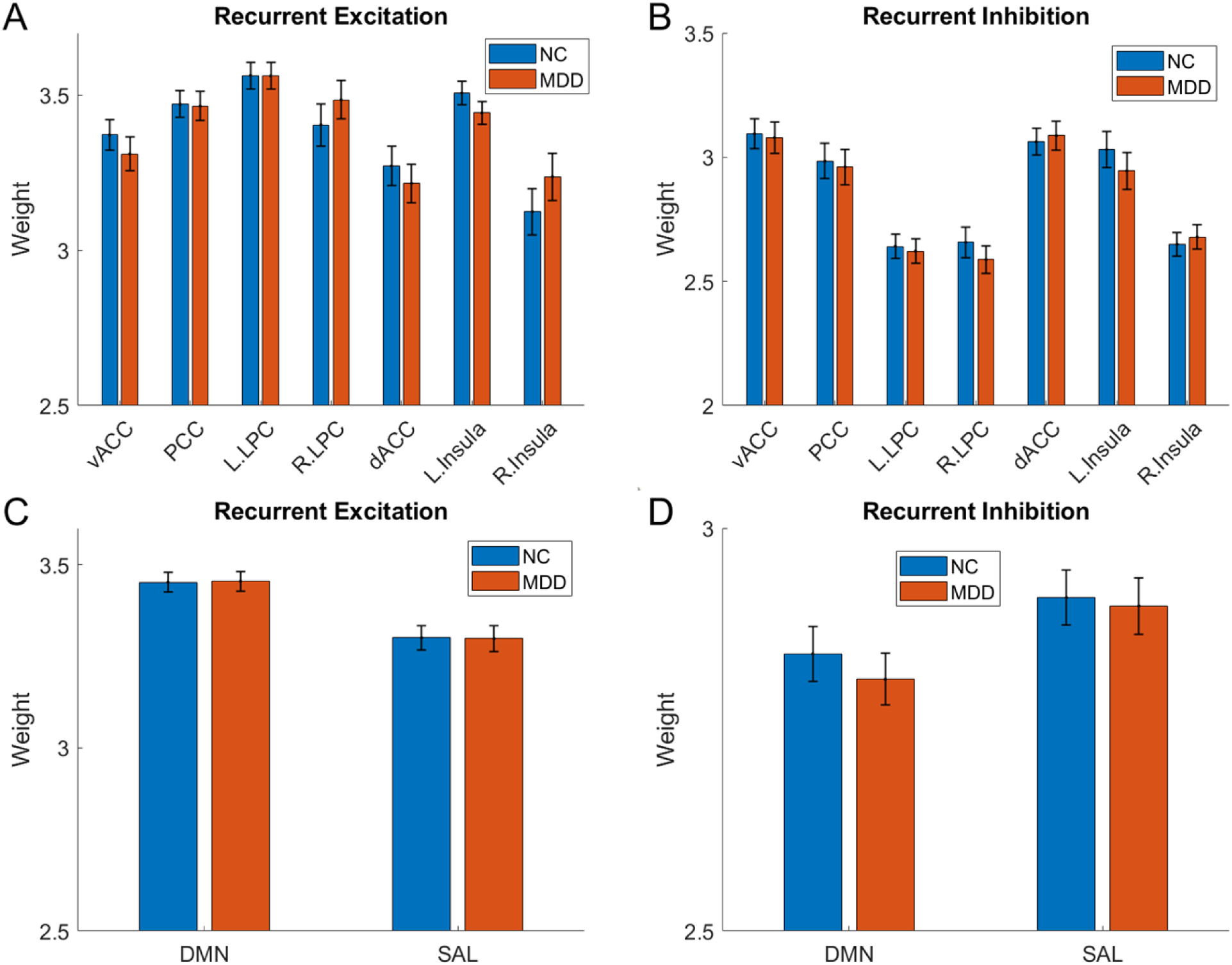
Average recurrent excitatory and inhibitory weight between NC and MDD populations in the DMN-SAL network. **(A)** Average recurrent excitatory weight within each ROI. **(B)** Average recurrent inhibitory weight within each ROI. **(C)** Average recurrent excitatory weight within the DMN and SAL networks. **(D)** Average recurrent inhibitory weight within the DMN and SAL networks. Error bars indicate standard error.

**Figure 7.**
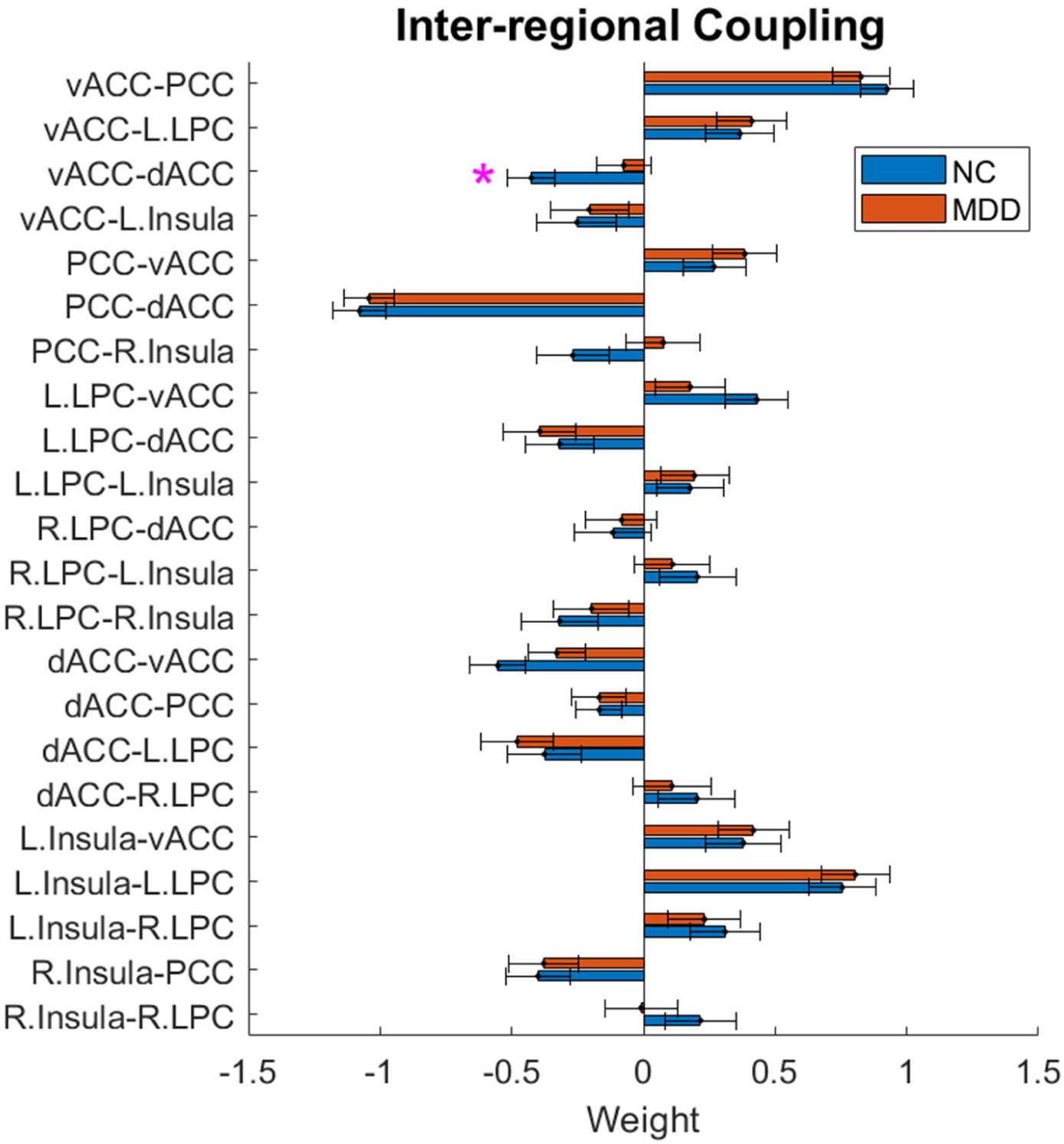
Average inter-regional EC between NC and MDD populations in the DMN-SAL network. The pink star indicates uncorrected significant connection (*p* < 0.05, uncorrected). Error bars indicate standard error.

### Significant EC group differences in the executive-limbic network

We next applied the MNMI approach to the executive-limbic network, another prevalence hypothetic model, and compared both intra- and inter-regional EC between the same two groups of subjects. The group average local recurrent excitation and inhibition weights and their network-specific averages are shown in Fig. 8. The inter-regional coupling weights are shown in Fig. 9. Different from the default mode-salience network, we observed a significant difference in the intra-regional connection weight. Besides a marginal increase for the MDDs in the recurrent excitation weight within the left SPC (*p* < 0.05, uncorrected; Fig. 8A), we found the recurrent inhibition weight within the left amygdala (in the limbic system) was significantly decreased in MDD (*p* < 0.05, FDR corrected; Fig. 8B). In addition, the network-averaged recurrent excitation weight within the EXE was significantly elevated in MDD (*p* < 0.05, FDR corrected; Fig. 8C), while the average recurrent inhibition weight within the LIM was significantly reduced in MDD (*p* < 0.05, FDR corrected; Fig. 8D) compared to NC.

**Figure 8.**
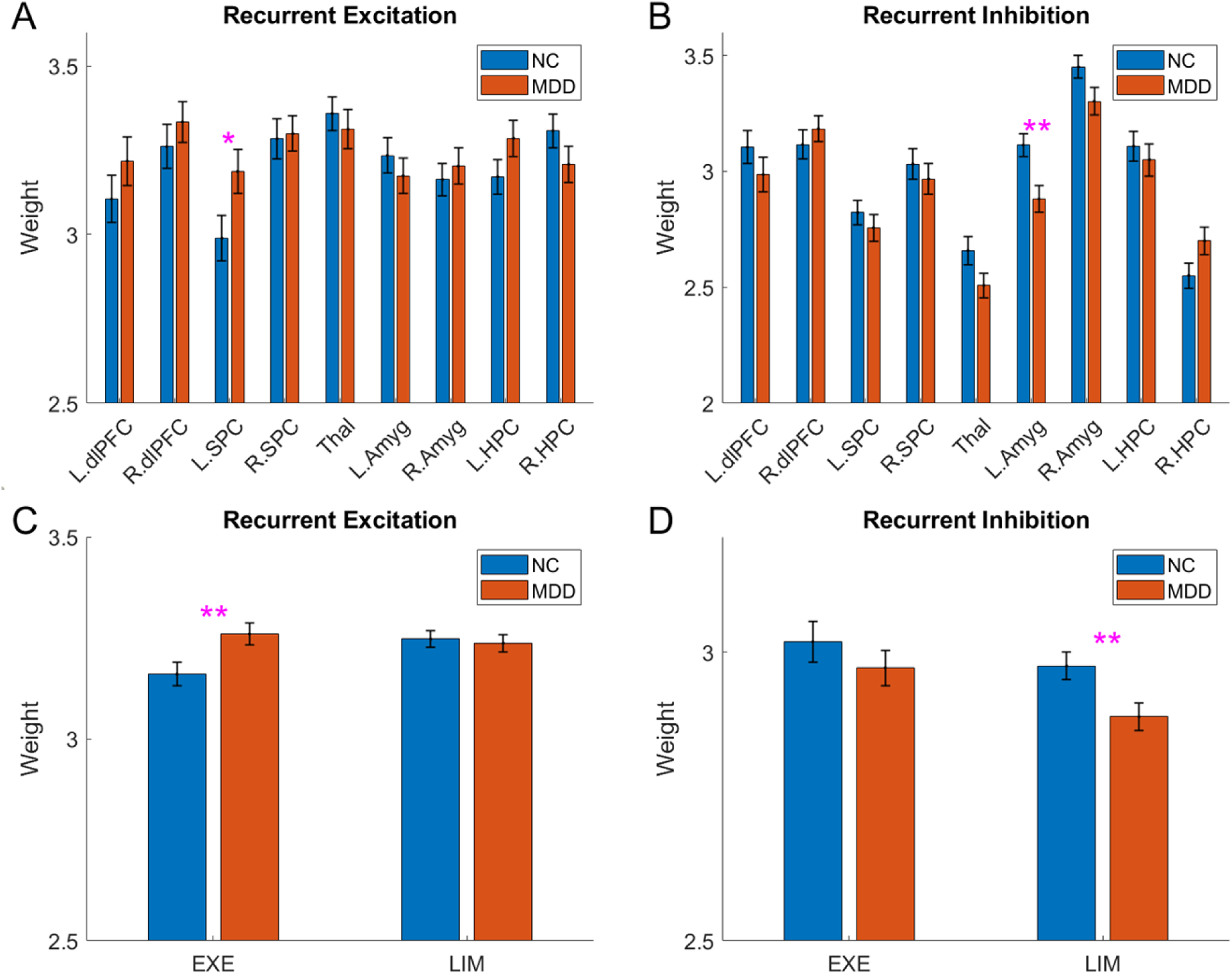
Average recurrent excitatory and inhibitory weight between NC and MDD populations in the EXE-LIM network. **(A)** Average recurrent excitatory weight within each ROI. **(B)** Average recurrent inhibitory weight within each ROI. **(C)** Average recurrent excitatory weight within the EXE and LIM networks. **(D)** Average recurrent inhibitory weight within the EXE and LIM networks. One pink star indicates uncorrected significant connections (*p* < 0.05, uncorrected), and double pink stars indicate corrected significant connections (*p* < 0.05, corrected by FDR). Error bars indicate standard error.

In addition to intra-regional EC, the inter-regional EC in the executive-limbic network also showed more significant differences in more connections between NC and MDD compared with the default mode-salience network (Fig. 9 and Fig. 10). In particular, the within-EXE connection from the left SPC to the right dlPFC was found to switch from excitatory (0.39) in NC to inhibitory (−0.18) in MDD (*p* < 0.05, FDR corrected) and the EXE-to-LIM connection from the left SPC to the right amygdala was significantly increased from 0.015 in NC to 0.56 in MDD (*p* < 0.05, FDR corrected). In addition, we observed that the EXE-to-LIM connection from the left dlPFC to the left hippocampus was significantly reduced (from 0.57 to 0.039; *p* < 0.05, FDR corrected) and its mirrored connection on the other hemisphere (from the right dlPFC to the right hippocampus) was marginally decreased (from 0.73 to 0.33; *p* < 0.05, uncorrected) in MDD. Moreover, the inhibitory EXE-to-LIM connection from the right SPC to the thalamus was found to be marginally reduced (from -0.47 to -0.12; *p* < 0.05, uncorrected). Nevertheless, we did not observe any significant difference in the network-averaged connection strength (Supplemental figure S7).

**Figure 9.**
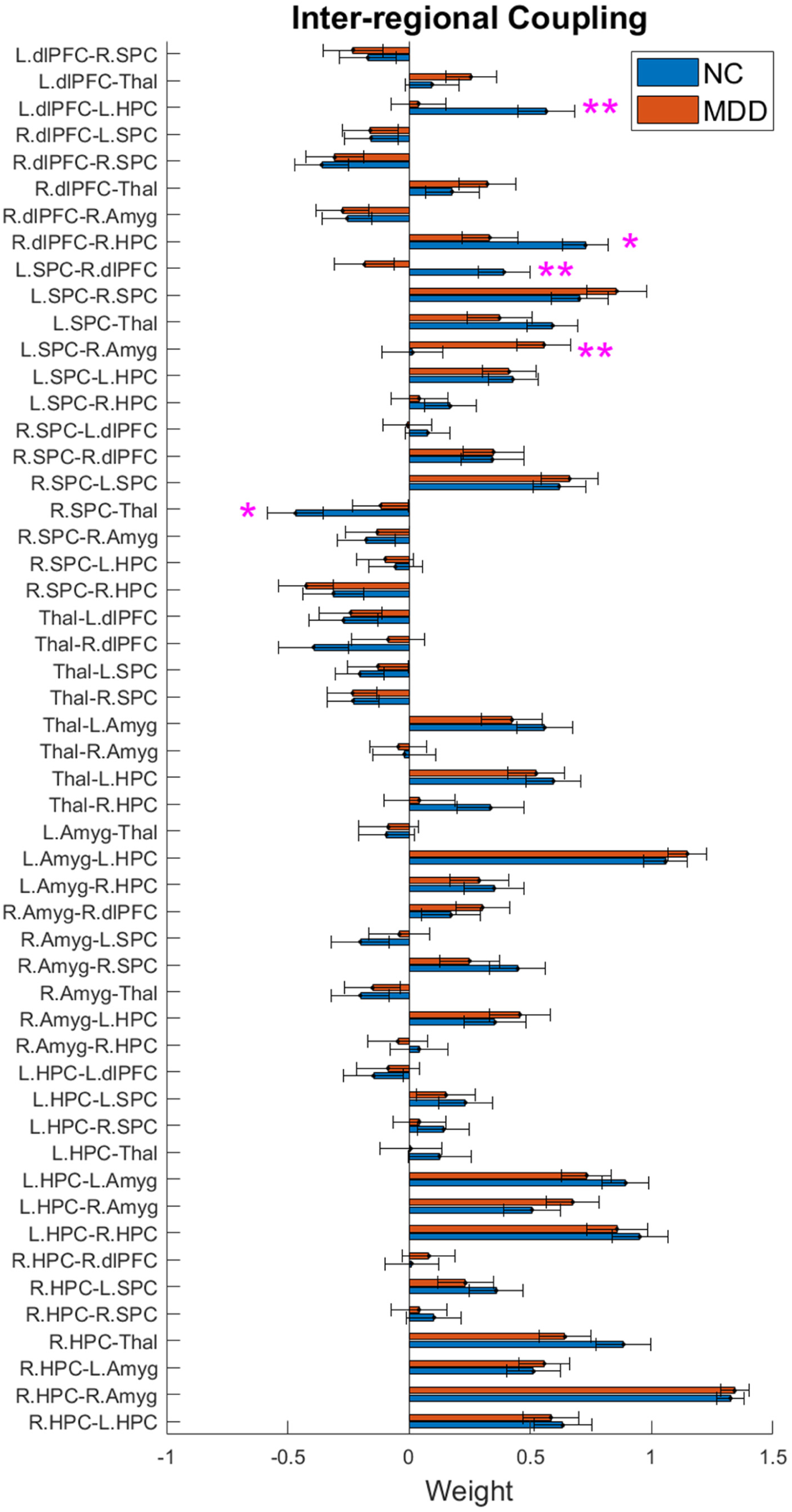
Average inter-regional EC between NC and MDD populations. One pink star indicates uncorrected significant connections (*p* < 0.05, uncorrected), and double pink stars indicate corrected significant connections (*p* < 0.05, corrected by FDR). Error bars indicate standard error.

**Figure 10.**
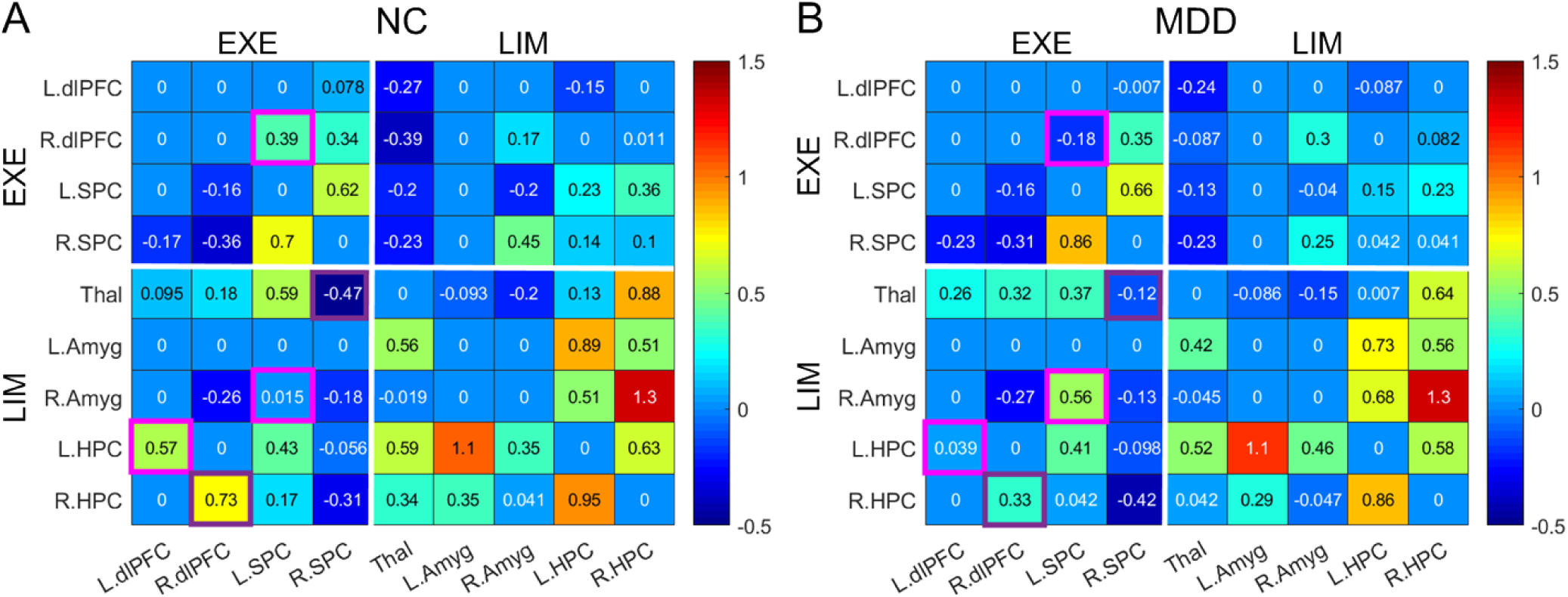
Connectivity matrices within the EXE-LIM network for NC and MDD groups. **(A)** Average inter-regional EC for the NC group. **(B)** Average inter-regional EC for the MDD group. The links with significant group difference are highlighted in pink (*p* < 0.05; FDR corrected) and purple (*p* < 0.05, uncorrected) boxes.

### A hypothetical model of executive-limbic malfunction in MDD

Based on our findings, we hypothesized that the executive-limbic network plays a central role in MDD pathophysiology and we thus proposed a hypothetical model that potentially explains the disrupted cortical-limbic interaction and dynamics leading to depressive symptoms (Fig. 11). According to this model, the recurrent excitation within the left SPC (in EXE) is substantially increased while the recurrent inhibition within the amygdala (in LIM) is greatly reduced in MDD, which could lead to over-excitation of these two brain regions. In the meantime, the EC from the left SPC to the right dlPFC switches from excitation in NC to inhibition in MDD, which might lead to diminished response in the right dlPFC. The SPC is involved in attending to perceptual cues in the environment (Vincent et al., 2008) and the dlPFC plays a pivotal role in attentional control and executive functioning (Elliott, 2003; Wang et al., 2018). Disrupted SPC and dlPFC activity may underlie biased attention for negative stimuli and impaired cognitive regulation of emotional processing in MDD (Fales et al., 2008; Beevers et al., 2010; Disner et al., 2011). In addition, as the EC from the right dlPFC to the right amygdala was inhibitory for both NC and MDD (Fig. 10A, B), the reduced right dlPFC response will decrease the inhibition on the right amygdala. Such a reduced inhibition, together with the significantly increased excitation from the left SPC to the right amygdala (due to increased left SPC activity and elevated L.SPC→RAmygdala connection strength), finally leads to overly elevated response of the right amygdala in MDD (Fig. 11). Because of the critical role of the amygdala in emotional processing and fear learning (LeDoux, 2000), overly excessive amygdala responses might lead to depressive symptoms including increased anxiety and cognitive bias over negative stimuli (Disner et al., 2011). In another pathway, the excitatory EC from the dlPFC (in EXE) to the hippocampus (in LIM) is significantly reduced in MDD, which could in turn decrease the hippocampal responses. Due to the important role of the hippocampus in memory function (Tulving and Markowitsch, 1998), reduced hippocampal excitation may be responsible for memory impairments (Hammar and Ardal, 2009) or biased memory for negative stimuli (Disner et al., 2011) in MDD. The impaired EXE-to-LIM EC also manifests as the decreased inhibition from the right SPC to the thalamus, which could in turn result in over-excitation of the thalamus. As the thalamus plays a central role in thalamocortical oscillations (Huguenard and McCormick, 2007; Li, et al., 2017), increased thalamic excitation may lead to impaired thalamocortical rhythms through its widespread projection to the neocortex, as observed in MDD patients (Llinas et al., 1999; Hughes and Crunelli, 2005). Since thalamocortical oscillations are associated with important brain functions including arousal, attention and sensory processing (Steriade et al., 1993; Buzsaki and Draguhn, 2004; Timofeev and Bazhenov, 2005), disrupted thalamocortical rhythms may lead to insomnia, anhedonia and impaired cognitive functions in MDD.

**Figure 11.**
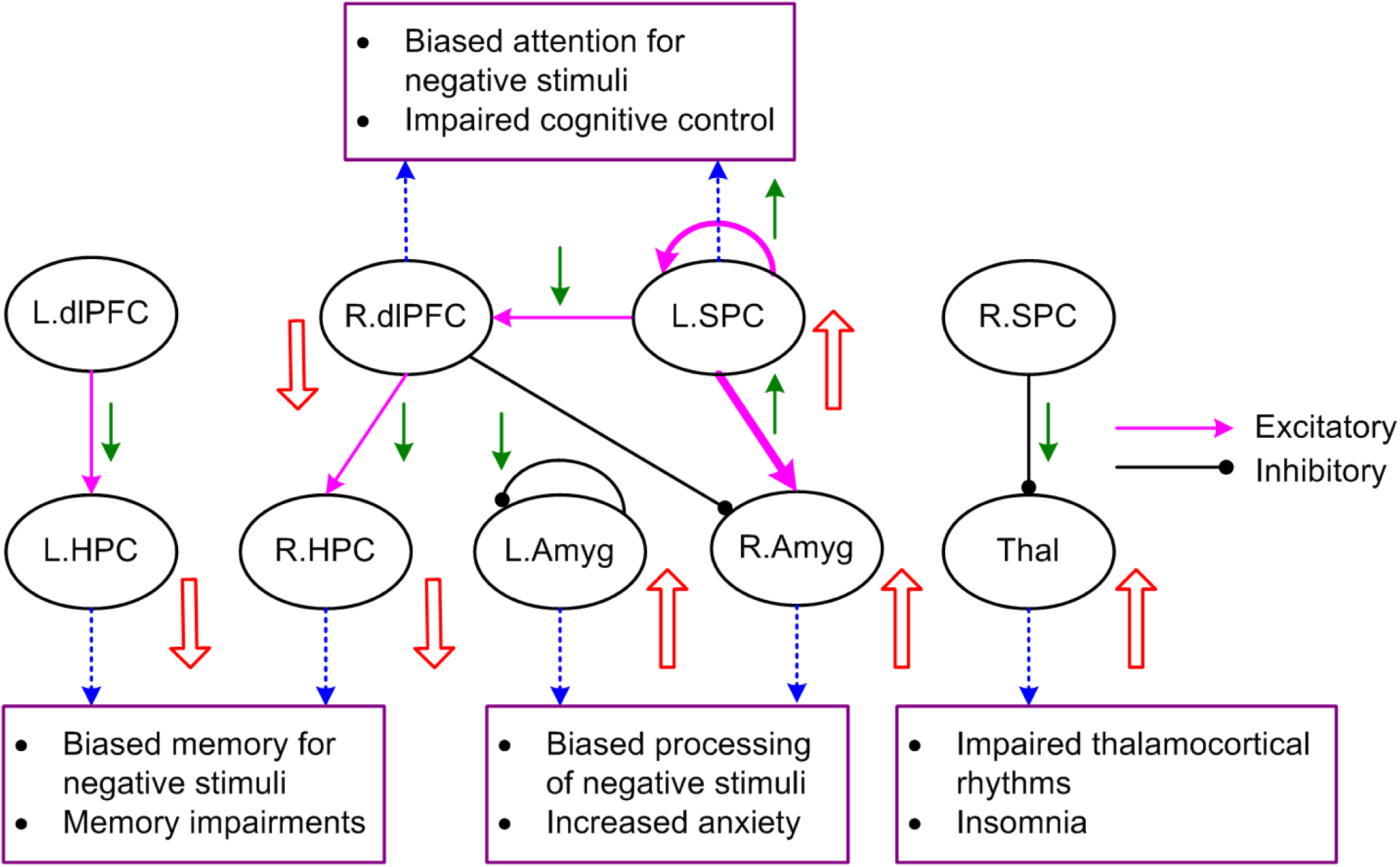
A hypothetical model of executive-limbic malfunction in MDD. MDD is mediated by increased recurrent excitation in the left superior parietal cortex (L.SPC) and reduced excitation (or increased inhibition) from the L.SPC to the right dorsolateral prefrontal cortex (R.dlPFC), leading to increased response of L.SPC and decreased response of R.dlPFC, which underlie biased attention for negative stimuli and impaired cognitive control function. Also, the recurrent inhibition within the left amygdala is deceased while the excitatory drive from the L.SPC to the right amygdala is significantly increased, resulting in amygdala hyperactivity that causes biased processing of negative stimuli and increased anxiety. In addition, the excitatory projection from the left/right dlPFC to the left/right hippocampus (HPC) is abnormally decreased, leading to reduced activity of the HPC underlying biased memory for negative stimuli and other memory impairments. Moreover, the inhibition from the R.SPC to the thalamus is reduced, which results in thalamic hyperactivity responsible for abnormal thalamocortical rhythms and insomnia. The green arrows indicate the change of the effective connectivity in MDD from normal control. The UP/DOWN arrows next to the brain regions indicate the change in neural responses in MDD compared to NC.

## Discussion

Major depressive disorder (MDD) is a complex mental disorder involving impairments in multiple brain networks and circuits as well as several neuromodulatory systems (Drevets et al., 2008; Price and Drevets, 2010; Pandya et al., 2012; Drysdale et al., 2017). Despite extensive research, the precise underlying mechanisms of MDD remain not well understood. Although previous functional neuroimaging studies have indicated hyper- or hypo-activity in multiple brain regions and/or interrupted functional connectivity (FC) among several large-scale functional systems (Price and Drevets, 2010; Disner et al., 2011; Menon, 2011; Dutta et al., 2014; Mulders et al., 2015), it is not well studied with fMRI (neither through task-related nor resting-state experimental protocol) how abnormal activity and/or connectivity are developed at multiple, e.g., both cellular and circuit, levels at the same time. Answering the questions which functional networks play a central role (origin of pathology) and which functional networks play a subordinate role (consequence of pathology) will not only provide deeper mechanistic insights of MDD pathophysiology, but also help to develop more effective treatments to target the pathological focuses of MDD. Towards this aim, we have developed a rs-fMRI-based, multiscale neural model inversion (MNMI) framework and tested its effectiveness on linking mesoscale cellular interactions intra-regionally with macroscale network connectivity inter-regionally in our previous proof-of-concept study (Li, et al., 2019). To make the model more realistic and enable the detectability of fine-grained abnormalities that could be easily overwhelmed by brute-force averaging for reducing freely estimable parameters, in this study, we substantially extended this framework toward computational efficiency and biological validity and applied it to test two further refined models regarding two competing hypotheses of MDD pathology. We demonstrated that MDD is more likely caused by disrupted interaction and dynamics in a core “executive-limbic” network rather than “default mode-salience” network, consistent with the limbic-cortical dysregulation model (Mayberg, 1997; Price and Drevets, 2010; Disner et al., 2011). Our improved MNMI approach went one step further and revealed detailed underlying mechanisms of executive-limbic dysregulation at the cellular and circuit levels.

### Inversion of a neural mass model based on rs-fMRI

Traditional computational analysis for fMRI data focuses on macroscopic connectome modeling such as undirected FC and graph theoretical analysis (Li K et al., 2009; Sporns, 2014). Despite their great success in characterizing the complex organizational topology of large-scale structural and functional brain networks, their clinical translation and application to fundamental neuroscience problems are still limited (Braun et al., 2018). One important reason is that such macroscopic network analysis is largely descriptive and cannot offer a mechanistic understanding of neural circuit function in healthy and diseased cohorts. Addressing this limitation requires a better integration of macroscopic connectome analysis with biophysically realistic neuronal models.

Dynamic causal modeling (DCM) is one of the neuroimaging computational methods to tackle this problem. It leverages a Bayesian framework to infer hidden neuronal mechanisms and architectures based on fMRI or electrophysiological data (Friston et al., 2003). By incorporating biologically plausible neural mass models, DCM of electrophysiological timeseries has addressed physiological questions related to human cognition (Moran et al., 2011), attention (Auksztulewicz and Friston, 2015), and cortical hierarchies (Bastos et al., 2015). By comparison, DCM for fMRI has primarily focused on a simplified state-space model (Friston et al, 2003), though two variations of DCM (stochastic DCM and spectral DCM) have been developed to specifically deal with rs-fMRI (Li B et al., 2011; Friston et al., 2014). To account for intrinsic (within region) connectivity, earlier DCM used a simple two-state model, extended from a one-state model, for each brain region (Marreiros et al., 2008) and it was not until recently the latest DCM for fMRI incorporated a more sophisticated, hierarchical neural mass model (Friston et al., 2019). Nevertheless, the latter DCM model currently applies to cortical circuits and task-based fMRI only. More recently, Wang et al., (2019) inverted a large-scale circuit model of the cerebral cortex using dynamic mean field modeling of rs-fMRI to study macroscale cortical hierarchy. Despite its applicability to rs-fMRI, it only contained a single type of neural population for each brain region and, in this model, the inter-regional connection weights were only allowed to vary homogenously for all connections, making it less realistic or plausible.

Our proposed MNMI model is directly applicable to rs-fMRI, incorporates realistic neural population dynamics, and enables independent estimation of individual EC links. Compared to DCM and its derivatives, our MNMI framework has several unique features. *First*, it uses the well-established Wilson-Cowan oscillator to model intrinsic neural dynamics due to its capability to capture large-scale neuronal interactions and global spontaneous activity during resting-state condition (Sejnowski, 1976, Amit and Brunel, 1997, Destexhe and Sejnowski, 2009; Abeysuriya et al., 2018). *Second*, the inter-regional connection weights are based on another direct connectivity measurement (structural connectivity from diffusion MRI). This introduced structural constraint on the neural mass model making it more realistic (Friston, 2011). We further removed weak links to construct sparse EC networks and improve computational efficiency. *Third*, we applied a biologically inspired genetic algorithm (GA) instead of the variational Bayesian scheme (i.e., expectation-maximization (EM) algorithm; Friston et al, 2003; Friston et al., 2007) to estimate EC. Compared with the EM algorithm, the GA does not require prior EC expectation and the number of free parameters is not limited by the number of unique entries in the FC matrix. Also, the GA allows easy computing parallelization when implemented in the MATLAB global optimization toolbox. Results indicated that the GA effectively found the optimal solution with quicker and more robust convergence performance. *Lastly*, our framework is highly flexible and can be easily customized. One can choose any specific type of neural mass model, the network architecture, the objective function, and the free parameters to estimate depending on the problem of interest. For example, one can model multiple layers within the cortical circuits to study layer-specific information processing.

### Executive-limbic malfunction as a core mechanism of MDD

One of the major finding of this study is that we identified executive-limbic malfunction as a core pathophysiological mechanism of MDD. It has long been suggested that it is limbic-cortical dysregulation that mediates the pathogenesis of MDD (Mayberg, 1997, 2003; Price and Drevets, 2010; Disner et al., 2011). However, for human MDD, it is unclear whether the dysregulation originates from limbic or cortical system and whether such dysregulation results from intrinsic (intra-regional) or inter-regional interactions. Our results demonstrated that both limbic and cortical systems and both intra-regional and inter-regional connectivities could play a role. *First*, we demonstrated that multiple intra-regional and inter-regional mechanisms underlie disrupted prefrontal-amygdala connectivity balance. Impaired prefrontal-amygdala interaction is a key component of the limbic-cortical dysregulation model (Price and Drevets, 2010; Disner et al., 2011) and an inverse relationship between amygdala and dlPFC activation has been observed (Fales et al. 2008), suggesting the existence of an inhibitory pathway from dlPFC to downregulate amygdala. Consistently, the model revealed an inhibitory EC from the right dlPFC to the right amygdala, for both NC and MDD (Fig. 10A, B). Also, the model suggests a critical role of the SPC in dlPFC hypoactivity and amygdala hyperactivity, and SPC affects dlPFC and amygdala activity through both intrinsic and external mechanisms. On one hand, the recurrent excitation within the left SPC was marginally increased in MDD, potentially leading to increased activity of this brain region. On the other hand, the EC from the left SPC to the right dlPFC switched from excitatory in NC to inhibitory in MDD and the excitatory EC from the left SPC to the right amygdala greatly increased, which may account for over-inhibition of the right dlPFC and over-excitation of the right amygdala (Fig. 11). Reduced activity of the right dlPFC would decrease inhibition on the right amygdala, further contributing to amygdala hyperactivity. It should be noted that the absolute EC from the left SPC to the right dlPFC was reduced in MDD (from 0.39 to -0.18; Fig. 10A, B), consistent with decreased frontoparietal connectivity in MDD revealed by a large-scale meta-analysis of resting-state FC (Kaiser et al., 2015). Moreover, the model indicated a critical role of intrinsic inhibition in amygdala hyperactivity, manifested by significantly reduced recurrent inhibition within the left amygdala (Fig. 8B). The amygdala is a heterogeneous structure consisting of multiple nucleus with excitatory and inhibitory intrinsic connections (Maren, 2001; Pare et al., 2004). The abnormally reduced recurrent inhibition within the left amygdala is consistent with the essential role of intrinsic inhibition in regulating amygdala activation and fear expression (Li et al., 2009, 2011).

Second, the model revealed disrupted prefrontal-hippocampus EC as an important contributor of the executive-limbic dysregulation. We observed that the excitatory projection from the dlPFC to the hippocampus was significantly reduced in MDD (Fig.10), potentially resulting in reduced activity in the hippocampus. This is in agreement with decreased hippocampal volume that has been consistently found in MDD (Otte et al., 2016), possibly because the prolonged stress leads to overall neuronal atrophy and synaptic depression in the hippocampus (Chaudhury et al., 2015; Thompson et al., 2015). The potentially reduced hippocampal excitation in MDD could be in opposite to the hyperactive amygdala. Such differential excitation/inhibition modulation agrees well with the experimental observations that unlike the hippocampus, the amygdala displays increases in volume in MDD (Frodl et al., 2003) as chronic stress could induce dendritic hypertrophy (Vyas et al., 2002).

Lastly, the model indicates impaired SPC-thalamus EC in the executive-limbic network. The thalamus is highly implicated in the MDD pathology (Drevets et al., 2008; Price and Drevets, 2010) as one previous study has also indicated impaired PFC-thalamus FC as a key feature of treatment-resistant depression (Li et al., 2013). We showed that the inhibitory EC from the right SPC to the thalamus was marginally reduced in MDD, which could potentially lead to elevated activity of the thalamus. Such a finding agrees with the experimental observations that during MDD, or the depressed phase of bipolar disorder, the glucose metabolism in the medial thalamus is abnormally increased (Price and Drevets, 2010). Thus, it is possible that the abnormal SPC-thalamus FC has already existed in early depression episode, which gives way to more significant PFC-thalamus disconnectivity in the more serious, treatment-resistant depression. It is noted that overall, the recurrent inhibition within the LIM system was significantly reduced, while the recurrent excitation within the executive network was substantially elevated (Fig. 8C, D). The decreased intrinsic inhibition in the limbic network mainly reflects reduced inhibition in the amygdala (Fig. 8B), while the increased recurrent excitation of the executive network mainly includes increased intrinsic excitation of the SPC (Fig. 8A).

### Role of default mode and salience networks in MDD

As mentioned earlier, many neuroimaging studies implicate the default mode and salience networks in the pathophysiology of MDD (Menon, 2011; Dutta et al., 2014; Mulders et al., 2015). Disrupted FC within the DMN has been frequently associated with excessive rumination and self-referential processing in MDD (Cooney et al., 2010; Hamiltonet al., 2015). However, in contrast to the executive-limbic network, the default mode-salience network only showed marginal difference in one inter-regional EC link (Fig. 7). The marginal difference in EC was not likely due to the excessive removal of weak connections because only two inter-regional connections displayed marginal significance in the network without removal of weak connections (Supplemental figure S6). Our findings suggest that the default mode-salience network may not be the core neural system underlying the pathogenesis of MDD. Instead, we think that they could rather constitute a secondary system whose dysfunction originates from the executive-limbic network (and then further induces FC or EC abnormalities in the DMN and SAL). Indeed, in an integrated cognitive-biological model of MDD, it is proposed that excessive ruminative thoughts and sustained self-referential processing associated with the DMN is facilitated by heightened emotional processing from the amygdala and sustained by attenuated top-down inhibition from the dlPFC and ventrolateral PFC (Disner et al., 2011). Subsequent disrupted connectivity in the DMN may become more stable as the disease progresses, as a recent large-cohort meta-analysis study reported reduced FC in the DMN only in recurrent MDD, but not in FEDN (Yan et al., 2019).

On the surface, our results may also be at odd with our previous study which shows that MDD is mainly associated with abnormal EC within the DMN and between the DMN and SAL networks (Li et al., 2020). Two main factors may account for such discrepancies. First, the analysis approach is different. In the previous study (Li et al. 2020), we used spectral DCM (Friston et al., 2014) to estimate inter-regional EC among the DMN, EXE, SAL and LIM networks, while in the current study, we used a newly developed MNMI framework to evaluate both intra- and inter-regional EC within the default mode-salience network and the executive-limbic network separately. The underlying neural model of spectral DCM is essentially a state-space model and it estimates inter-regional EC in the *frequency* domain (Friston et al., 2014). By comparison, the MNMI method used the more biologically realistic neural mass model and estimated both intra-regional and inter-regional EC in the *time* domain. The increased biological realism may enable us to reveal more fundamental neurophysiological mechanisms of depression. Second, as mentioned above, dysfunctional connectivity within the default mode-salience network may be caused by executive-limbic malfunction. When the default mode-salience network is isolated from the executive-limbic network, most of the abnormal connectivity patterns within the default mode-salience network may disappear. It should be noted that in Li et al. (2020), we did observe substantial abnormal EC links involving the executive and limbic networks (Fig. 5B of Li et al., 2020).

### Clinical implication to MDD treatment

Our findings indicate executive-limbic malfunction as a core mechanism of MDD pathophysiology. Specifically, the model predicts elevated neural activity in the superior parietal cortex, amygdala, and thalamus, along with reduced neural activities in the dlPFC and hippocampus. Based on such a hypothetic mechanism, effective treatments can be proposed to restore normal activity patterns in these brain regions to alleviate depressive symptoms. Indeed, repetitive transcranial magnetic stimulation (rTMS), an effective therapeutic modality for MDD, predominantly targets the dlPFC. The stimulation presumably increases dlPFC’s excitability with concomitant downstream effects on the limbic system (Li et al., 2004; Fox et al., 2012; Leuchter et al., 2013). According to our model, boosted dlPFC activity would impose stronger inhibition on the amygdala, reducing biased processing of emotional stimuli. Besides, deep brain stimulation (DBS) of the inferior thalamic peduncle (ITP) has been shown to be effective to treatment-resistant MDD (Raymaekers et al., 2017; Dandekar et al., 2018). The ITP is a bundle of fibers that reciprocally interconnect with the thalamic reticular nucleus (TRN; Drobisz and Damborska, 2019), a subcortical structure that exerts strong inhibition on the dorsal thalamic nuclei (Pinault, 2004). Thus, stimulation of the ITP most likely reduces thalamic excitability via the TRN inhibition, which is well aligned with our model. Our model also suggests that stimulation that can dampen amygdala and superior parietal cortex activity while enhancing hippocampus excitability may have therapeutic benefit for MDD.

## Conclusions

Using a multiscale neural model inversion (MNMI) framework, we demonstrated that major depressive disorder is more likely characterized by disrupted circuit interactions within the executive-limbic network, rather than the default mode-salience network. We found that the impaired frontoparietal effective connectivity within the executive network may contribute to hypoactivity in dlPFC, while decreased intrinsic inhibition combined with increased SPC excitation could lead to amygdala hyperactivity, together resulting in predominant PFC-amygdala activation imbalance in MDD. In addition, we observed disrupted top-down PFC-hippocampus and PPC-thalamus connectivity in MDD that could contribute to impaired memory function and abnormal thalamocortical oscillations. Our findings support the long-standing notion that limbic-cortical dysregulation underlies the pathogenesis of MDD. Future treatments should specifically target the executive-limbic system for maximal therapeutic benefits.

## Materials and Methods

### Subjects

We used the same fMRI dataset as our previous study (Li et al., 2020) except excluding two NCs and four MDDs who have excessive head motion (mean frame-wise displacement (FD) (Power et al., 2012) is more than three time the standard deviation from the mean of their respective population), resulting in 98 NC and 96 MDD subjects (note that in a previous proof-of-concept MNMI study (Li et al., 2019), we included 66 NC and 66 MDD subjects). The demographic and clinical characteristics of the participants are shown in Table 1. The FEDN patients were recruited from the psychological counseling outpatient of the First Affiliated Hospital of Guangzhou University of Chinese Medicine, Guangdong, China from September 2015 to June 2018. The MDD diagnosis was carried out by two experienced professional psychologists according to the 17-item Hamilton Rating Scale for Depression (HDRS-17; Hamilton, 1967) and the Diagnostic and Statistical Manual 5th edition (DSM-5; American Psychiatric Association, 2013). The selected FEDN patients were right-handed native Chinese speakers aged between 18 and 55 years old who were firstly diagnosed with MDD and had no history of any neurological illness or any other forms of psychiatric disorders. The healthy subjects were enrolled locally during the same period of time and were physically and mentally healthy based on their medical history and the Mini-International Neuropsychiatric Interview (Sheehan et al., 1998) with a total HDRS-17 score of less than seven. The study was approved by the local ethics committee and all participants provided written informed consent complying with the Declaration of Helsinki.

### FMRI acquisition and preprocessing

The MRI acquisition was based on a 3.0-T GE Signa HDxt scanner with an 8-channel head-coil within three days of diagnosis and the preprocessing procedures were detailed in Li et al. (2020) and briefly described as follows. The acquisition parameters for the rs-fMRI were: TR (repetition time) = 2000 ms, TE (echo time) = 30 ms, flip angle = 90°, matrix size = 64 × 64, and slice spacing = 1.0 mm, and those for the structural MRI were: slice thickness = 1.0 mm (156 slices), no slice gap, matrix = 256 × 256, field of view = 256 × 256 mm^2^. Image preprocessing was performed using SPM12 (www.fil.ion.ucl.ac.uk/spm) and Data Processing Assistant for Resting-State fMRI (DPARSF) version 2.3 (http://rfmri.org/DPARSF) (Yan and Zang, 2010). For each subject, 180 rs-fMRI volumes were remained after removing the first five volumes. The remaining images were corrected for slice acquisition timing and head motion. The structural MRI was used to guide rs-fMRI registration by using unified segment and Diffeomorphic Anatomical Registration through Exponentiated Lie Algebra (DARTEL). The rs-fMRI data was smoothed with a 6-mm full-width-at-half-maximum (FWHM) Gaussian kernel and denoised by regressing out several nuisance signals, including the Friston-24 head motion parameters and signals from cerebrospinal fluid and white matter, followed by linear detrending and temporal band-pass filtering (0.01-0.08 Hz).

### Structural connectivity

To introduce structural constraint on EC, structural connectivity was assessed based on probabilistic tractography with the diffusion MRI data from the Human Connectome Project. Data from 14 unrelated and randomly selected subjects was processed and the resulting structural connectivity matrices were averaged. Specifically, for each of the 14 subjects, tissue segmentation was performed on the structural MRI (aligned with the diffusion MRI data) from the 14 subjects based on FreeSurfer (https://surfer.nmr.mgh.harvard.edu/) according to the pipeline described in (Smith et al., 2012). After minimal preprocessing of the diffusion MRI data, probabilistic tractography was employed in the diffusion MRI’s native space using the first-order integration over fiber orientation distributions (iFOD1) algorithm estimated by spherical deconvolution approach that considers fiber crossing (Tournier et al., 2010) to reconstruct two million streamlines within the whole brain with random seeds. The output streamlines were cropped at the grey matter-white matter interface with a search distance of 2 mm, where the Destrieux atlas (included with FreeSurfer) that contains 164 regions of interest (ROIs) (Destrieux et al., 2010) was applied, resulting in a 164 × 164 structural connectivity matrix. Each element is the number of streamlines between a pair of ROIs normalized by the average volume of them, representing the structural connectivity between them. Out of the full structural connectivity matrix, we selected (see next section) a partial structural connectivity matrix of the default mode-salience network and that of the executive-limbic network, separately. They were further normalized by the maximal entry of the two matrices so the structural connectivity was bounded between 0 and 1.

### Functional connectivity

From the Destrieux atlas, we selected 16 representative ROIs (Table 2, visualized in Supplemental figures S1-S4) in each of the four functional networks (DMN, SAL, EXE and LIM) based on their presumptive important roles in MDD neuropathology (Price and Drevets, 2010; Menon, 2011; Dutta et al., 2014; Mulders et al., 2015; Drysdale et al., 2017). Of note, more representative ROIs could be easily included as a natural extension of our method in the future. The default mode-salience network included seven ROIs: ventral anterior cingulate cortex (vACC, combining both left and right portion of it since it is in the medial part of the brain, similarly for other midline brain regions unless specified otherwise), posterior cingulate cortex (PCC, both ventral and dorsal parts), and left and right lateral parietal cortices (LPC, each side represents one ROI) from DMN, as well as dorsal anterior cingulate cortex (dACC), and left and right insula from SAL. The executive-limbic network contained nine ROIs: left and right dorsolateral prefrontal cortices (dlPFC), and left and right superior parietal cortices (SPC) from EXE, as well as the thalamus, left and right amygdala, and left and right hippocampus from LIM. Since the brain parcellation was carried out by Freesurfer in each subject’s native space, to apply those ROIs to the preprocessed rs-fMRI data in the MNI space, we used *FNIRT* in FSL (https://fsl.fmrib.ox.ac.uk/fsl) to estimate the deformation field based on each subject’s structural MRI after it was aligned to the same subject’s rs-fMRI, which generated the transformation from the native structural MRI space to the standard MNI space. This transformation was then used to warp the partitions from the native structural MRI space to the rs-fMRI space. Regional averaged BOLD rs-fMRI time series were extracted from the 16 ROIs and, the FC matrix was calculated using Pearson’s correlation. To estimate more accurate FC by removing the effect of potential noise and artifacts, we divided the total 360-s BOLD time series into ten 180-s sliding windows with an interval of 20 s. The final FC was computed by averaging the ten corresponding FC matrices after removing the outliers (more than three scaled median absolute deviations away from the median) for each link.

**Table 2.**
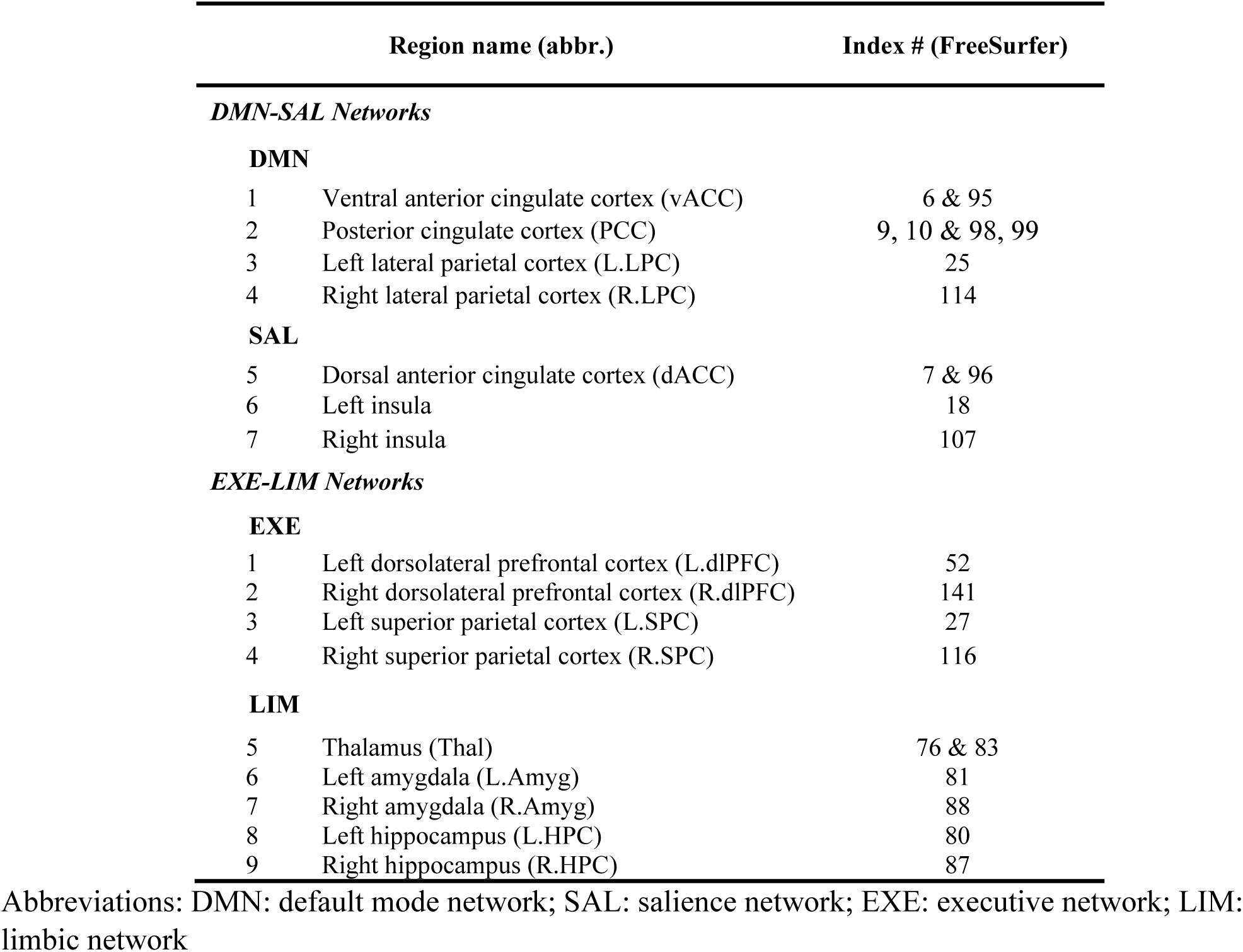
ROI definitions of the default mode-salience and executive-limbic networks with corresponding index numbers in the Destrieux atlas included in FreeSurfer

### Network modeling of neural activity

We applied computational neuronal modeling to capture neural interactions both within and between brain regions. Each brain region consists of reciprocally coupled excitatory and inhibitory neural populations (Fig. 1). The regional brain dynamics is simulated by a neural mass model using the biologically motivated nonlinear Wilson-Cowan oscillator (Wilson and Cowan, 1972). The population-level activity of the j^th^ region is governed by the following two equations (Abeysuriya et al., 2018):

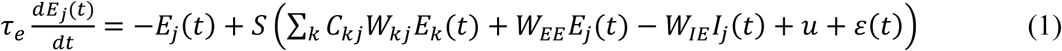

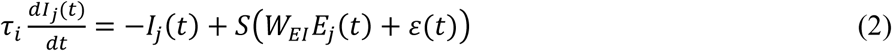

where *E_j_* and *I_j_* are the mean firing rates of the excitatory and inhibitory neural populations in brain region *j*, τ*_e_* and τ*_i_* are the excitatory and inhibitory time constants (20 ms; Hellyer et al., 2016), *W_EE_*, *W_EI_* and *W_IE_* are the local connection strengths from excitatory to excitatory neural population, from excitatory to inhibitory neural population and from inhibitory to excitatory neural population, respectively, *u* is a constant external input, and *ε(t*) is random additive noise following a normal distribution centering at 0 with standard deviation of 0.3. It is important to mention that the constant input *u* and noisy input *ε(t*) represent the extrinsic inputs from other un-modeled brain regions to allow the generalization ability of the model. The long-range connectivity strength from region *k* to region *j* is represented by *W_kj_* (and that from region *j* to *k* is *W_jk_*) that is derived from empirical structural connectivity and scaled by an inter-regional coupling factor *C_kj_* to model individual difference. The nonlinear response function *S* is modeled as a sigmoid function 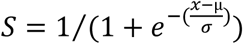 (μ=1.0; σ =0.25; Abeysuriya et al., 2018).

### Hemodynamic modeling

The neural activity of each brain region is converted into corresponding BOLD signal using a well-established hemodynamic model (Friston et al. 2003). Specifically, for each region *j*, the fluctuation in neuronal activity *x_j_ (E_j_* and *I_j_*) gives rise to a vasodilatory signal *s_j_* that is subject to self-regulation. The vasodilatory signal causes change in the blood flow *f_j_* leading to subsequent change in blood volume *v_j_* and deoxyhemoglobin content *q_j_*. The hemodynamic state equations with parameters are given by Friston et al. (2003):

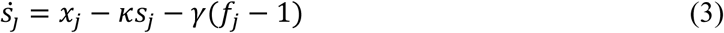

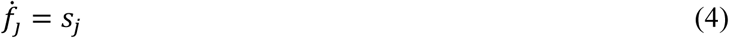

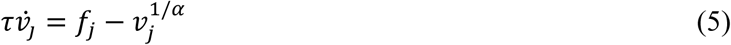

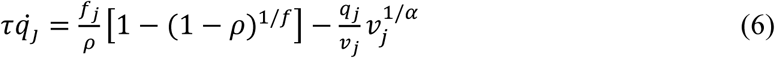

where *κ* is the rate of decay (0.65 per second), *γ* is the rate of flow-dependent elimination (0.41 per second), *τ* is the hemodynamic transit time (0.98 s), *α* is the Grubb’s exponent (0.32) and *ρ* is the resting oxygen extraction (0.34). The neural activity from excitatory and inhibitory populations was summed up within each region with respective weighting (2/3 for excitatory and 1/3 for inhibitory; Becker et al., 2015) to obtain the overall regional neural activity *x_j_*. The simulated BOLD signal is taken to be a static nonlinear function of volume and deoxyhemoglobin that depend on the relative contribution of intravascular and extravascular components:

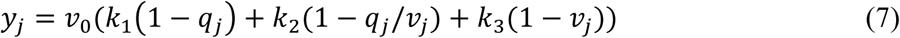

where *ν*_0_ is the resting blood volume fraction (0.02), and *k*_1_*, k*_2_ and *k*_3_ are the intravascular, concentration and extravascular coefficients, respectively (*k*_1_ *=* 7*ρ, k*_2_ = 2, and *k*_3_ *= 2ρ* − 0.2).

### Numerical integration

The Wilson-Cowan and hemodynamic models were simulated using the 4^th^ order Runge-Kutta (RK) scheme with an integration step of 10 ms (Wang et al., 2019; shorter integration step had no significant effect on the results reported). We simulated the network for a total of 200 s, and the first 20 s of the BOLD activity was discarded to remove transient effects. The remaining 180-s (3-min) time series were downsampled to 0.5 Hz to have the same temporal resolution as the real, recorded BOLD rs-MRI signals.

### Estimation of model parameters

The free parameters in the neural network model include both local (intra-regional) and long-range (inter-regional) ECs. For the local parameters, we estimated both the recurrent excitation weight (*W_EE_*, excitatory to excitatory) and recurrent inhibition weight (*W_IE_*, inhibitory to excitatory) in each brain region. The excitatory to inhibitory weight (*W_EI_*) was assumed to be constant and equaled to three (Abeysuriya et al., 2018); as the effect of *W_EI_* could be accommodated by the changes in *W_IE_*. The inter-regional coupling factor *C_kj_* was also estimated in the neural mass model. As mentioned, the default mode-salience network contained seven ROIs and the executive-limbic network consisted of nine ROIs, which gave rise to 42 and 72 inter-regional connections, respectively (self-connections were excluded). To improve model estimation efficiency, we performed a one sample *t*-test on each structural connection link across the 14 subjects from the Human Connectome Project and only preserved the links with strong structural connectivity (*p* < 0.0001, uncorrected). We did this selection because we aimed to focus on only strong (thus most putative) direct connections and EC depends on structural connectivity (Friston, 2011). Twenty links were removed from each of the default mode-salience and executive-limbic networks, resulting in 22 and 52 inter-regional connections for the two much sparser networks, respectively. In addition, the external input (*u*) was also estimated and assumed to be identical for all brain regions of a subject. Consequently, a total of 37 (14 local, 22 inter-regional, and one for shared external input) parameters were estimated for the default mode-salience network and a total of 71 (18 local, 52 inter-regional, and one for shared external input) parameters were estimated for the executive-limbic network.

We used the genetic algorithm (a function *“ga ”*) contained in the MATLAB’s global optimization toolbox to estimate the model parameters. The genetic algorithm is a biologically inspired method for solving both constrained and unconstrained optimization problems based on natural selection, the process that drives biological evolution (Mitchell, 1995). The genetic algorithm repeatedly modifies a population of individual solutions. At each step, it selects individuals at random from the current population to be parents and uses them to produce the children for the next generation. Over successive generations, the population "evolves" toward an optimal solution. The genetic algorithm differs from other regular optimization algorithms in that it generates a population of points (instead of a single point) at each iteration and uses stochastic (instead of deterministic) computation to select the next population, which is suitable for optimization problems involving discontinuous, non-differentiable, stochastic or highly nonlinear objective functions. In the MNMI framework, the parameters were bounded within certain ranges to achieve balanced excitation and inhibition in the network. Based on previous studies (Abeysuriya et al., 2018), we assumed a default value of 3.0 for both recurrent excitation (*W_EE_*) and inhibition (*W_IE_*) strength, and a default value of 0.3 for the external input (*u*). Therefore, we set the searching ranges around the default values, i.e., [2.0 4.0] for both recurrent excitation and inhibition weights, and [0.2 0.4] for the external input, by allowing a maximum of 33.3% variation from the default value. We allowed the inter-regional connection weights to be either positive (excitatory) or negative (inhibitory), so we set the searching range to be [-2 2] for the inter-regional coupling factor (*C_kj_*). This allows a relatively wide range for inter-regional EC weights, but at the same time, avoids unconstrained parameter estimation that could generate extreme and unrealistic estimates. Note that we assumed the inter-regional inhibition could occur directly between excitatory neural populations (instead of via local inhibitory neurons) for simplicity.

We set the objective function to be the distance, measured by the opposite of the Pearson’s correlation, between the simulated and empirical FC matrices, so that the genetic algorithm could maximize the Pearson’s correlation between the simulated and empirical FC for each subject. The tolerance was set to be 0.001 and the maximal number of generations was set to be 128. We observed good convergence within 128 generations for all the subjects and an even smaller tolerance did not change the estimated parameters in any significant way. The genetic algorithm repeatedly generated a population of solutions (or parameter set) based on stochastic computation in each iteration and evaluated the objective function by numerically integrating the neural mass and hemodynamic models (for each parameter set) until the average change in the fitness value (the value of the objective function) was less than the functional tolerance. The entire MNMI optimization procedure with model integration was coded with MATLAB (R2018b) and run at a Linux-based high-performance cluster. The genetic algorithm program was run in parallel with 12 cores using the Parallel Computing Toolbox in MATLAB. The typical computing time (for each individual subject) ranged from 10 to 20 hours for the default mode-salience network and 12 to 30 hours for the executive-limbic network with a few subjects taking more than 30 hours.

### Statistical analysis

The model parameters were estimated for each subject and compared between the NC and MDD groups. We used a two-sample *t*-test to compare each of the estimated model parameters and the significant level was set to a false discovery rate (FDR) of *q* < 0.05. We did not use the network-based statistics (NBS; Zalesky et al., 2010) as our previous paper (Li et al., 2020) because we estimated both intra- and inter-regional EC and we removed weak inter-regional connections making it difficult to fulfill the topological cluster requirement of NBS.

### Data and code availability

The in-house developed MATLAB code that support the findings of this study are available from the GitHub repository at https://github.com/Guoshi-Li/MNMI_MDD. The structural and diffusion MRI data for the 14 subjects used to construct structural connectivity was randomly selected from the WU-Minn HCP Data (1200 subjects) available from the Connectome Database (https://db.humanconnectome.org/). The raw fMRI data is not publicly available according to the ethic committee of the First Affiliated Hospital of Guangzhou University of Chinese Medicine but can be shared upon reasonable requirement directed to the imaging PI (Dr. Shijun Qiu).

## Data Availability

The in-house developed MATLAB code that support the findings of this study are available from the GitHub repository at http://github.com/Guoshi-Li/MNMI_MDD. The structural and diffusion MRI data for the 14 subjects used to construct structural connectivity was randomly selected from the WU-Minn HCP Data (1200 subjects) available from the Connectome Database (http://db.humanconnectome.org/). The raw fMRI data is not publicly available according to the ethic committee of the First Affiliated Hospital of Guangzhou University of Chinese Medicine but can be shared upon reasonable requirement directed to the imaging PI (Dr. Shijun Qiu).

## Acknowledgements

G.L., Y.W. and P.-T.Y. were supported by an NIH grant (EB022880). H.Z. and D.S. were supported by NIH grants (EB022880, AG042599, AG049371, and AG041721). H.Z. was also supported by an NIH grant (MH108560). Y.L., Y.Z., and S.Q. were supported by National Natural Science Foundation of China - Major International (Regional) Joint Research Project (81920108019), Major Project (91649117), and General Project (81771344, and 81471251), as well as Innovation and Strong School Project of Education Department of Guangdong Province (2014GKXM034), and Science and Technology Plan Project of Guangzhou (2018–1002-SF-0442). Y.L. was also supported by China Scholarship Council (201708440259) and Excellent Doctoral and PhD Thesis Research Papers Project of Guangzhou University of Chinese Medicine (A1-AFD018181A55). D.L. was supported by Traditional Chinese Medicine Bureau of Guangdong Province (20202059).

